# Metabolomics pilot study identifies circadian desynchronization and distinct intra-patient variability patterns in critical illness

**DOI:** 10.1101/2019.12.09.19014225

**Authors:** Elizabeth R. Lusczek, Lee Parsons, Jesse Elder, Stephen B. Harvey, Mariya Skube, Sydne Muratore, Greg Beilman, Germaine Cornelissen-Guillaume

## Abstract

**Background:** Synchronized circadian rhythms play a key role in coordinating physiologic health. Desynchronized circadian rhythms may predispose individuals to disease or be indicative of underlying disease. Intensive care unit (ICU) patients likely experience desynchronized circadian rhythms due to disruptive environmental conditions in the ICU and underlying pathophysiology. This observational pilot study was undertaken to determine if circadian rhythms are altered in ICU patients relative to healthy controls by profiling circadian rhythms in vital signs and plasma metabolites.

**Methods:** We monitored circadian rhythms in 5 healthy controls and 5 ICU patients for 24 hours. Heart rate and blood pressure were measured every 30 minutes, temperature was measured every hour, and blood was sampled for mass spectrometry-based plasma metabolomics every 4 hours. Bedside sound levels were measured every minute. Circadian rhythms were evaluated in vitals and plasma metabolites individually and in each group using the cosinor method.

**Results:** ICU patient rooms were significantly louder than healthy controls’ rooms and average noise levels were above EPA recommendations. Healthy controls generally had significant circadian rhythms individually and as a group. While a few ICU patients had significant circadian rhythms in isolated variables, no significant rhythms were identified in ICU patients as a group, except in cortisol. This indicates a lack of coherence in circadian phases and amplitudes among ICU patients. Finally, principal component analysis of metabolic profiles showed surprising patterns in plasma sample clustering. Each ICU patient’s samples were clearly discernable in individual clusters, separate from a single cluster of healthy controls.

**Conclusions:** ICU patients’ circadian rhythms show significant desynchronization compared to healthy controls. Clustering of plasma metabolic profiles suggests that metabolomics could be used to track individual patients’ clinical courses longitudinally. Our results show global disordering of metabolism and the circadian system in ICU patients which should be characterized further in order to determine implications for patient care.

## Introduction

Circadian rhythms span multiple levels of hierarchical organization in biological systems, through molecular processes (peripheral oscillators) to whole-body rhythms such as core body temperature (1). Synchronized, intact rhythms are fundamental to human health and their desynchronization is now known to contribute to morbidity and pathophysiology in otherwise healthy individuals (2-4). Critical care physicians increasingly recognize the influence of circadian desynchronization on physiology and outcomes in critically ill patients (5, 6). There is clear evidence that supporting circadian rhythms in cancer patients contributes to higher quality of life (7). Likewise, it may be beneficial to support circadian rhythms in intensive care unit (ICU) patients (8, 9).

While there is evidence that circadian rhythms are disrupted in the critically ill, the etiology is complex and multifaceted (10). A host of desynchronizing influences are present in critical illness, including poor sleep (11), continuously-administered parenteral nutrition (12-14), unregulated light/dark cycles and sound levels (15-17), as well as the influence of pharmacological treatments (18).

Due to the hierarchical and widely distributed organization of the circadian system, multiple data types should be collected to determine the extent of circadian desynchronization in critical illness regardless of the underlying cause. High-throughput –omics data, particularly metabolomics, is suited to this endeavor, since metabolism is tightly coupled to the core clock mechanism (19-21). Multiple circadian metabolites have been reported in the literature, identified with mass spectrometry-based metabolomics (22-24) and alterations to circadian rhythms in the metabolome have been documented in shift work (25) and sleep disruption protocols (26).

We collected preliminary data to evaluate the extent of circadian desynchronization in ICU patients using mass spectrometry-based metabolomics in this observational study. Secondary objectives were to evaluate circadian desynchronization in vital signs and to measure sound levels in patient rooms. We hypothesized that disruptions to circadian rhythms in ICU patients relative to healthy controls would be identified in multiple metabolites and in vital signs. To test this hypothesis, we conducted a pilot study in 5 healthy controls and 5 ICU patients. This study is the first to use metabolomics to profile circadian rhythms in the critically ill.

## Methods

The study protocol (study number 1505M70361) was reviewed and approved by the University of Minnesota Institutional Review Board in accordance with the Code of Federal Regulations, 45 CFR 46.101(b). All enrolled healthy participants gave their informed consent to study staff. Informed consent was obtained from legally authorized representatives of all enrolled ICU patients since they were sedated and unable to consent.

### Patient population

From 2015-2016, circadian rhythms were monitored in 5 healthy control participants and 5 intensive care unit (ICU) patients for 24 hours. Healthy participants were over 18 years of age with a self-reported average of 7-8 hours of sleep a night, and were in generally good health. They were screened to exclude the following factors that may affect normal circadian rhythms: shift work, blindness, use of over-the-counter sleep aids or supplements, history of traumatic brain injury, brain surgery, or sleep apnea, smoking, use of psychotropic drugs, or current alcohol use of more than 2 drinks per day. Healthy participants were monitored by trained staff at the University of Minnesota’s Masonic Clinical Research Unit. Catheters were placed peripherally to facilitate blood draws. They were not subjected to constant routine conditions. They stayed in single rooms with windows allowing natural light. They were provided meals from a cafeteria at times of their choosing that generally corresponded to set mealtimes, controlled the room’s light levels to their preference, and determined their own periods of rest and activity. All 5 participants reported that they slept well despite nighttime monitoring.

ICU patients were recruited from the University of Minnesota Medical Center ICU according to the following criteria: age > 18 years, mechanical ventilation with sedation for at least 3 days, and any of the following: emergent surgery, emergent intubation, admission after severe trauma, damage control surgery, or admission to ICU for post-operative complication. Patients with these conditions were excluded: severe nervous system disorder such as traumatic brain injury or recent neurosurgery, confirmed or suspected drug overdose or alcohol abuse, blindness, severe renal failure, end stage liver disease or hepatic encephalopathy, extracorporeal membrane oxygenation (ECMO) use, required vasopressors or steroids, central sleep apnea, hemoglobin levels ≤8 g/dL, and dementia or delirium prior to ICU stay. All ICU patients were fed enterally.

Twelve participants were enrolled in the study. One healthy control was disqualified because of abnormally high blood pressure. One ICU patient was extubated during the study and disqualified.

### Data collection

In all 10 research participants, beginning at 9:00 AM, heart rate and blood pressure were measured every 30 minutes. Temperature was measured every hour orally (healthy controls) or via Foley catheter (ICU patients). Bedside sound levels were measured every minute (Bruel and Kjaer 2250L, Duluth, GA).

For the metabolomics portion of the study, blood samples (5 mL) were drawn for metabolomics analysis every 4 hours beginning at 9:00 AM in each participant. Plasma was extracted and aliquots were stored at −80°C until preparation for analysis with mass spectrometry. Reagents were obtained from Fisher Chemical Co., and were of LC-MS grade or better. Samples were prepared according to published protocols (27, 28). Internal standards (see S1 Table) were added to thawed plasma samples followed by the addition of 4 volumes of cold (−80°C) 10% acetone 90% methanol (i.e. added 400µL solvent to 100µl sample). Samples were vortexed and incubated at −20°C for 15 minutes. Samples were spun down at 13,000g for 10 minutes at 4°C and the supernatant transferred to a clean tube. The incubation and centrifugation were repeated and the final supernatant transferred to a clean tube, which was then dried under nitrogen. Preparation for reverse-phase LC-MS analysis was completed by adding a starting buffer (100 µl of 5% acetonitrile, 95% water, 0.1% formic acid). Sample pH was adjusted to approximately 2 by adding 10-20 µl of 10% formic acid.

Samples were analyzed via chromatographic separation in-line with mass spectrometry. Ultra-high performance liquid chromatography (UHPLC) was performed using the Thermo Scientific Ultimate 3000 UHPLC platform. For reverse-phase analysis the instrument was fitted with a Waters Acquity BEH C-18 column (2.1×100 mm, 1.7 µm particle size). Flow rate was 0.4 mL/minute and the column compartment was set to 40°C. Ten microliters were injected onto the column. Elution solutions were A) water with 0.1% formic acid and B) acetonitrile with 0.1% formic acid. The elution gradient was as follows: 2% B for 0.5 minutes; increase to 25% B over the next 0.5 minutes; increase to 80% B over the next 7 minutes; increase to 100% B for 2 minutes; decrease to 2% B over the next 0.5 minutes; hold at 2% B for 2.5 minutes.

The Q Exactive™ Quadrupole-Orbitrap mass spectrometer (Thermo Fisher Scientific, Waltham, MA) was employed for mass analysis. The instrument profiles for both high mass accuracy (<2 ppm) and spectral resolution (typically at 70,000 resolution in full scan mode). Analysis was performed in positive mode over a mass range of 70-1050 m/z.

### Data analysis

#### Evaluation of sound levels

Differences in bedside sound levels in participants’ rooms were evaluated using Wilcoxon Rank Sum tests with the R software package v.3.3.3 (29). Sound meter data were discarded for three study subjects (one healthy control and two ICU patients) due to technical failure. There was very little variation in the sound levels recorded for the three participants with flat noise levels (mean LAeq=49.0 dB, standard deviation=7.03 dB for analyzed participants; mean LAeq=25.7 dB, standard deviation=0.184 dB for discarded participants).

#### Evaluation of 24-hour rhythms

Time series measurements of each participant’s heart rate, blood pressure, temperature, and plasma metabolite intensities were evaluated for 24-hour rhythmicity using the cosinor methodology (30) in individuals and at the population level. MESOR (midline estimating statistic of rhythm or rhythm-adjusted mean), amplitude, and acrophase were determined for each variable in each individual. Individual p-values were combined in each group using the methodology described in Sokal and Rohlf (31). Briefly, the test consists of computing −2 * ∑*_k_ ln* (*P_k_*) where *k* is the number of p-values being combined (in this case, 5) and *P*_k_ are the individual p-values. This quantity follows a chi-square distribution with 2*k* degrees of freedom and a chi-square test is used to arrive at a final p-value indicative of whether or not individuals in each group had a significant circadian rhythm. This approach simplifies the evaluation and presentation of the individual rhythms, otherwise 150 p-values would need to be presented.

Population means for each variable were evaluated and compared between the two study groups using parameter tests for the population mean cosinor (PMC) (30). Inference is done by (1) calculating the arithmetic mean of individual MESORs and (2) calculating the average of individual amplitude/acrophase pairs, treating the pairs as vectors. PMC p-values indicate the statistical significance of the zero-amplitude test and reflect similar amplitude-acrophase pairs among individuals in the population at the trial period considered (i.e., 24 hours), irrespective of whether a circadian rhythm could be demonstrated individually. Note that PMC can show a significant result when the combined p-values do not because the PMC algorithm uses all the data at once, increasing the statistical power over the individual calculations.

#### Mass spectrometry metabolomics

Plasma metabolites were putatively identified with the Progenesis QI (Nonlinear Dynamics, Durham, NC) and Xcalibur (Thermo Fisher Scientific, Waltham, MA) software packages in conjunction with the Metlin (32) and HMDB (33) databases and an in-house database compiled by the facility performing the spectrometry. A group of 60 previously identified circadian metabolites were targeted (24, 34). Principal component analysis of all features identified with Progenesis QI was performed using the R software package after intensities were log-transformed (29).

Features were filtered using the ANOVA algorithm in Progenesis (q-value<0.05) when comparing ICU patients to healthy controls (35). Filtering resulted in a list of 15,000 identified features distinguishing ICU patients from healthy controls. This reduced dataset was linked with HMDB identifiers by Progenesis QI and exported directly to Ingenuity Pathway Analysis (QIAGEN Inc., https://www.qiagenbioinformatics.com/products/ingenuity-pathway-analysis/). IPA was able to map 12,825 of these HMDB IDs to its database. For features that matched more than one HMDB identity, IPA resolves duplicates by choosing the feature with the lowest p-value. Following the resolution of duplicates, the resulting dataset of 2,696 metabolites with HMDB IDs was used by IPA in the pathway analysis.

Serum cortisol, quantified by Fairview Hospital Laboratories, was used to confirm cortisol quantified by mass spectrometry.

## Results

Healthy controls (two males and three females) were 45-72 years of age. ICU patients (two males and three females) were 43-66 years of age. Patient characteristics are described in Table 1. BMI data for both groups is shown in Supporting Information S2, and did not differ between the groups.

**Table 1:** Pilot study ICU patient characteristics.

| Patient ID | APACHE II | Discharge Destination | Primary Diagnosis | Length of Stay |
| --- | --- | --- | --- | --- |
| P07 | 6 | Home | Tracheal stenosis | 8 days |
| P08 | 11 | Home | Tricuspid valve regurgitation | 30 days |
| P10 | 9 | Long-Term Acute Care | Necrotizing pneumonia, bronchopleural fistula | 23 days |
| P11 | 23 | Long-Term Acute Care | ARDS of unknown origin | 28 days |
| P12 | 14 | Long-Term Acute Care | Abscess requiring emergent tracheostomy | 23 days |

ICU patient demographics, APACHE II scores, and outcomes. ARDS=Acute Respiratory Distress Syndrome; APACHE=Acute Physiology and Chronic Health Evaluation

Sound levels were significantly higher in ICU patients’ rooms than in healthy controls’ rooms, particularly at night (Fig 1). Average minimum equivalent continuous noise levels (LAeq) were 48 dB (ICU) vs. 33 dB (controls; p<0.0001). Sound levels in ICU patient rooms were, on average, above recommended World Health Organization limits of 35 dB (36). In two of the patients’ rooms, the minimum sound levels were quite loud: 43.5 and 54.9 dB, respectively. Mean noise levels overlapped in ICU patient rooms and healthy controls’ rooms from 9:00 to 13:30 and again at 9:00 the following morning. Noise levels remained elevated in ICU patients’ rooms even during nighttime hours while healthy controls had a clear decrease in sound levels at night, particularly around midnight. We noted a clear lack of a 24-hour pattern in ICU noise levels relative to healthy controls.

**Fig 1:**
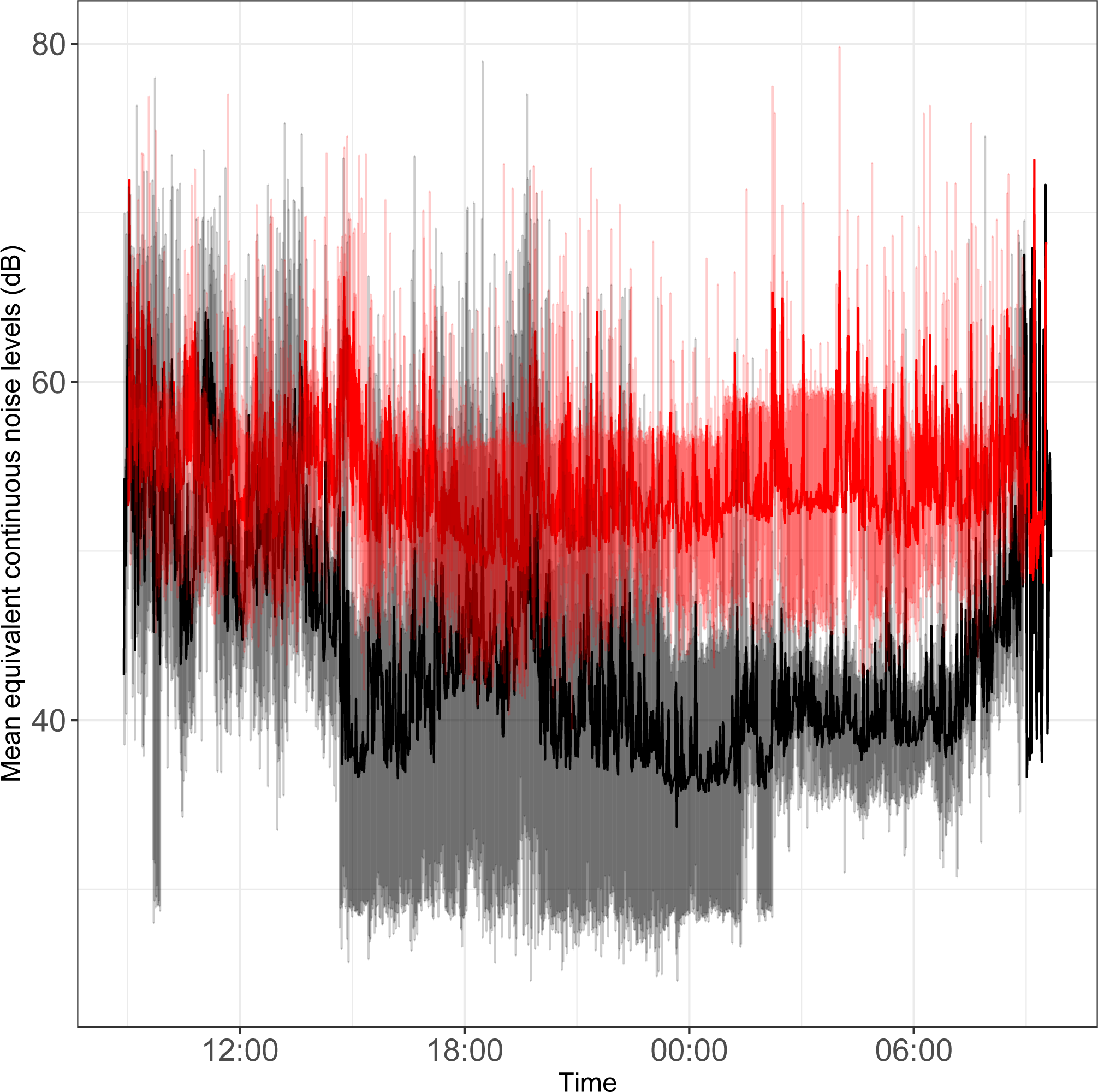
Time series plot of the mean equivalent continuous noise levels. Bedside noise levels were recorded in the rooms of ICU patients (red line) and healthy controls (black line). Light red shading represents the standard deviation for ICU patients; gray shading represents the standard deviation for healthy controls.

### 24-hour rhythms in vitals

24-hour rhythms exist both individually and at the group level in heart rate and blood pressure for healthy controls. Isolated individual ICU patients showed significant circadian rhythms in vital signs. Combined individual p-values, reflecting the degree to which rhythms were observed in individual study participants, are shown in Table 2. Low combined p-values reflect that significant rhythms were observed in multiple individuals. For comparison, PMC p-values are shown as well. The presence of significant rhythms in vitals at both the individual and population levels in healthy controls shows that these individuals had similar amplitudes and acrophases.

**Table 2:**
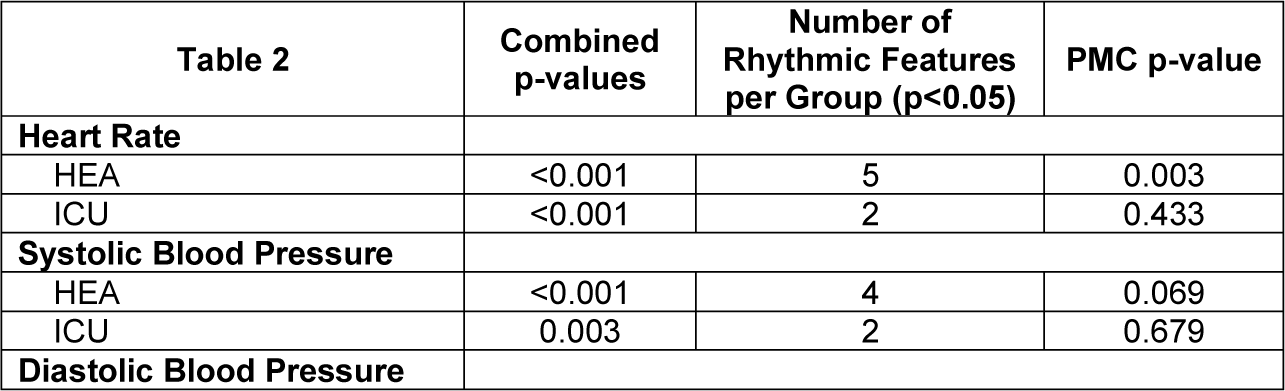
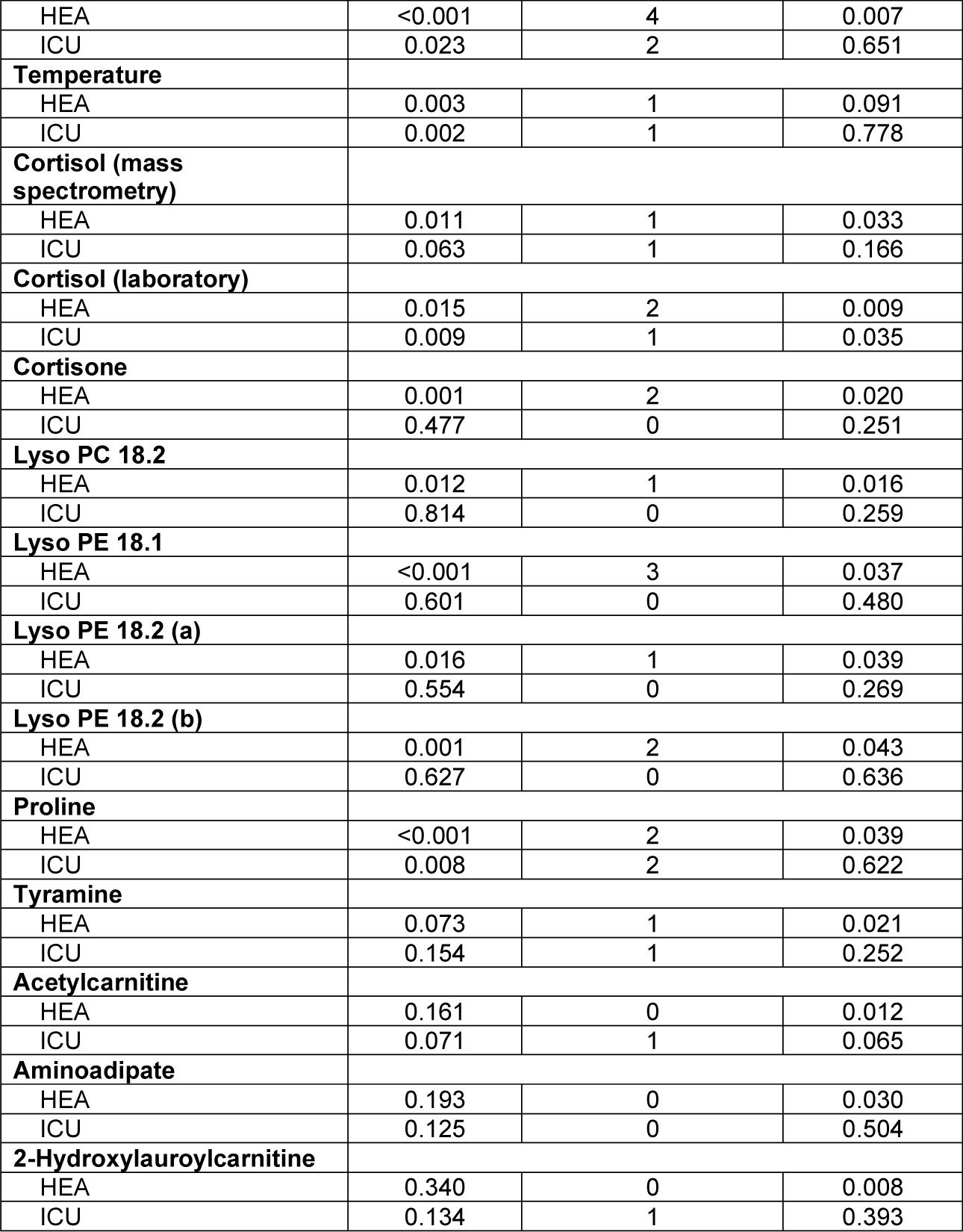
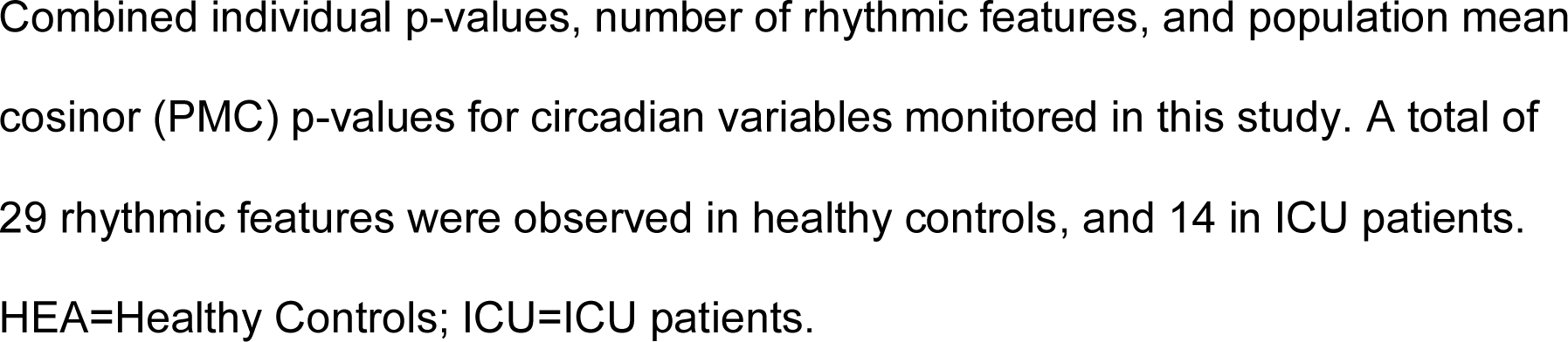
Significant circadian vitals and putatively identified metabolites.

| Table 2 | Combined p-values | Number of Rhythmic Features per Group (p<0.05) | PMC p-value |
| --- | --- | --- | --- |
| **Heart Rate** |  |  |  |
| HEA | <0.001 | 5 | 0.003 |
| ICU | <0.001 | 2 | 0.433 |
| **Systolic Blood Pressure** |  |  |  |
| HEA | <0.001 | 4 | 0.069 |
| ICU | 0.003 | 2 | 0.679 |
| **Diastolic Blood Pressure** |  |  |  |
| HEA | <0.001 | 4 | 0.007 |
| ICU | 0.023 | 2 | 0.651 |
| **Temperature** |  |  |  |
| HEA | 0.003 | 1 | 0.091 |
| ICU | 0.002 | 1 | 0.778 |
| **Cortisol (mass spectrometry)** |  |  |  |
| HEA | 0.011 | 1 | 0.033 |
| ICU | 0.063 | 1 | 0.166 |
| **Cortisol (laboratory)** |  |  |  |
| HEA | 0.015 | 2 | 0.009 |
| ICU | 0.009 | 1 | 0.035 |
| **Cortisone** |  |  |  |
| HEA | 0.001 | 2 | 0.020 |
| ICU | 0.477 | 0 | 0.251 |
| **Lyso PC 18.2** |  |  |  |
| HEA | 0.012 | 1 | 0.016 |
| ICU | 0.814 | 0 | 0.259 |
| **Lyso PE 18.1** |  |  |  |
| HEA | <0.001 | 3 | 0.037 |
| ICU | 0.601 | 0 | 0.480 |
| **Lyso PE 18.2 (a)** |  |  |  |
| HEA | 0.016 | 1 | 0.039 |
| ICU | 0.554 | 0 | 0.269 |
| **Lyso PE 18.2 (b)** |  |  |  |
| HEA | 0.001 | 2 | 0.043 |
| ICU | 0.627 | 0 | 0.636 |
| **Proline** |  |  |  |
| HEA | <0.001 | 2 | 0.039 |
| ICU | 0.008 | 2 | 0.622 |
| **Tyramine** |  |  |  |
| HEA | 0.073 | 1 | 0.021 |
| ICU | 0.154 | 1 | 0.252 |
| **Acetylcarnitine** |  |  |  |
| HEA | 0.161 | 0 | 0.012 |
| ICU | 0.071 | 1 | 0.065 |
| **Aminoadipate** |  |  |  |
| HEA | 0.193 | 0 | 0.030 |
| ICU | 0.125 | 0 | 0.504 |
| **2-Hydroxylauroylcarnitine** |  |  |  |
| HEA | 0.340 | 0 | 0.008 |
| ICU | 0.134 | 1 | 0.393 |

Combined individual p-values, number of rhythmic features, and population mean cosinor (PMC) p-values for circadian variables monitored in this study. A total of 29 rhythmic features were observed in healthy controls, and 14 in ICU patients. HEA=Healthy Controls; ICU=ICU patients.

As a group, healthy controls had a significant (PMC p<0.007) 24-hour rhythm in heart rate and diastolic blood pressure, and near-significant (PMC p<0.091) rhythms in temperature and systolic blood pressure (Fig 2 and Table 2). As a group, ICU patients had no significant 24-hour rhythms in the same variables, even though some patients had a significant rhythm at the individual level. This indicates a lack of coherence in the acrophases and amplitudes of ICU patients in these variables. This can also be seen in Table 3, which reports 24-hour rhythm characteristics (24-hour p-value, MESOR, amplitude, and acrophase) determined by PMC. For ICU patients, 95% confidence limits could not be computed for the majority of the variables measured. 24-hour rhythm characteristics of vital signs and putatively identified metabolites quantified with mass spectrometry, quantified with PMC. MESOR is the midline estimating statistic of rhythm, amplitude is the circadian amplitude, and acrophase reflects the timing of overall high values recurring each day, expressed in clock time. Confidence intervals for insignificant 24-hour rhythms are reported as NS (not significant). Mass spectrometry features are uniquely identified by retention time (RT) and mass to charge ratio (m/z) or neutral mass.

**Table 3:**
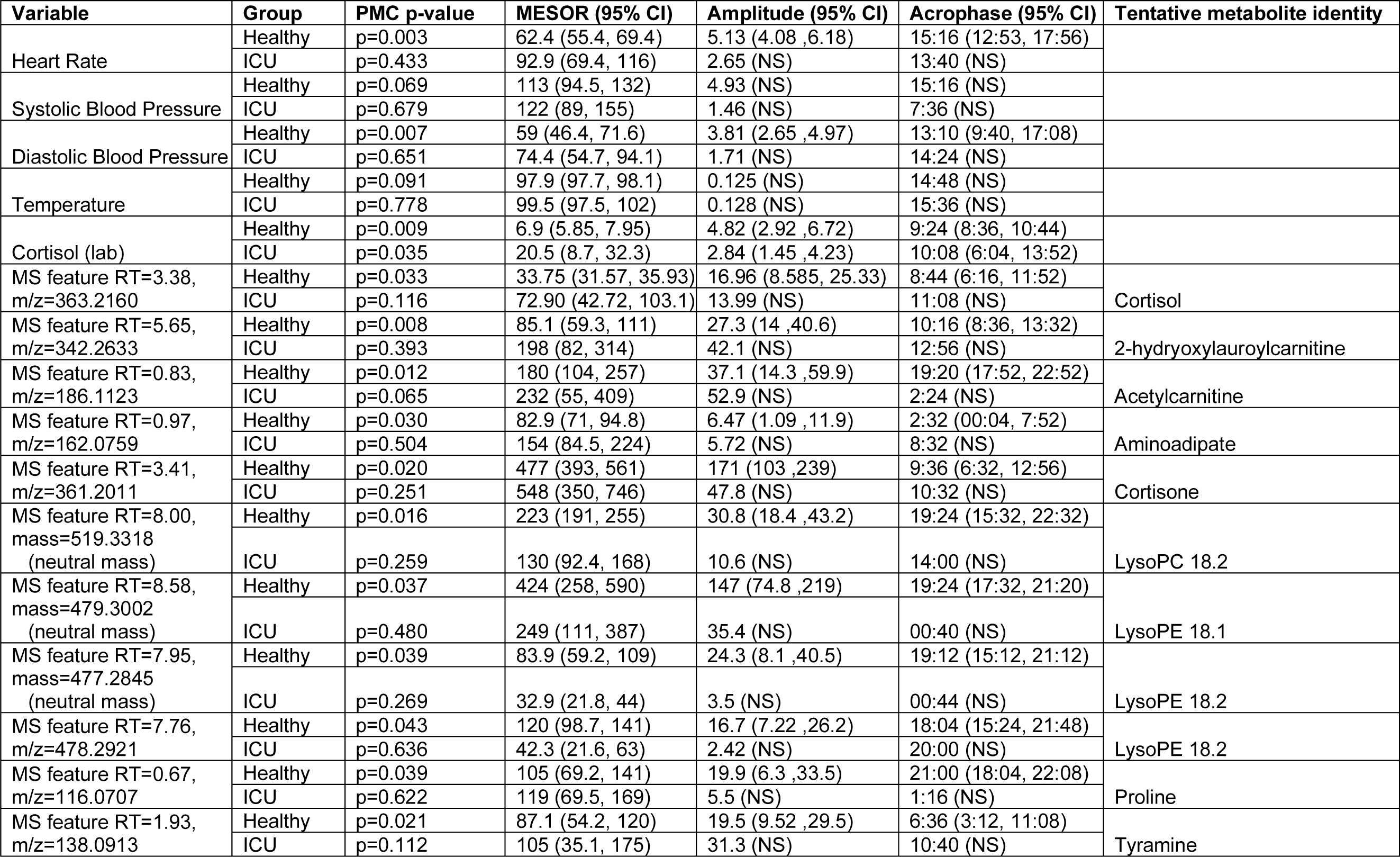
PMC Characteristics of vital signs and putatively identified metabolites.

**Fig 2:**
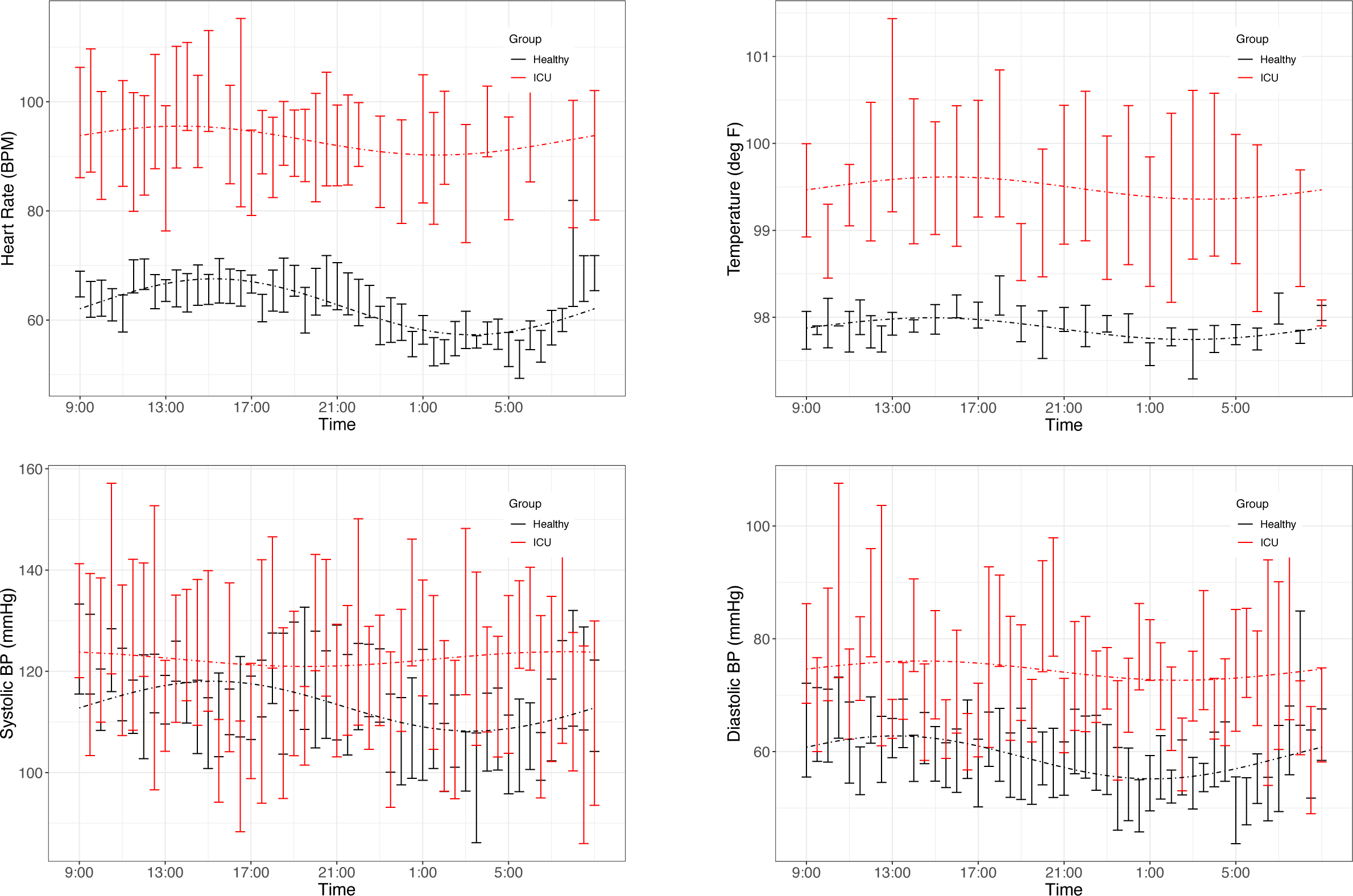
Time series plots of vitals. Data are presented as mean and standard error in ICU patients (red) and healthy controls (black). Clockwise: heart rate, temperature, diastolic blood pressure, systolic blood pressure. Dashed lines represent the PMC fits of the 24-hour rhythm to the data. Plots of the individual data are presented in S3 Fig.

### 24-hour rhythms in the metabolome

Progenesis QI identified 25,000 unique features. Sixty circadian metabolites were putatively identified using retention times and mass-to-charge ratios published in the literature (24, 34), online databases (METLIN, HMDB) and in-house lists curated by the Center for Mass Spectrometry and Proteomics. Of these 60 metabolites, PMC showed that 10 had a significant 24-hour rhythm in the healthy control group (Table 3 and S3 and S4 Figs) and none had a significant rhythm in the ICU group. For brevity, we present information only on the 10 significant metabolites.

Significant 24-hour rhythms were identified in cortisol both individually and at the group level in healthy controls and in ICU patients (Laboratory value, Table 2). Even though PMC showed that both the ICU group and the healthy control group had a significant 24-hour rhythm in cortisol, patients’ cortisol rhythms had some abnormal features: they had a higher mean (20.5 vs. 6.9 μg/dL, p=0.01) than controls as well as scattered acrophases (Fig 3). Healthy controls had tightly clustered acrophases (9:24 (8:36, 10:44)) while ICU patients did not (10:08 (6:04, 13:52); data are reported as acrophase (95% confidence interval)). The scatter of phases in the ICU patient group leads to a significant lowering of rhythm amplitude in the ICU patient group relative to the healthy control group as quantified by PMC. (Cortisol was quantified by the hospital laboratory and with mass spectrometry, showing good agreement in patterns (see S5 Fig)).

**Fig 3:**
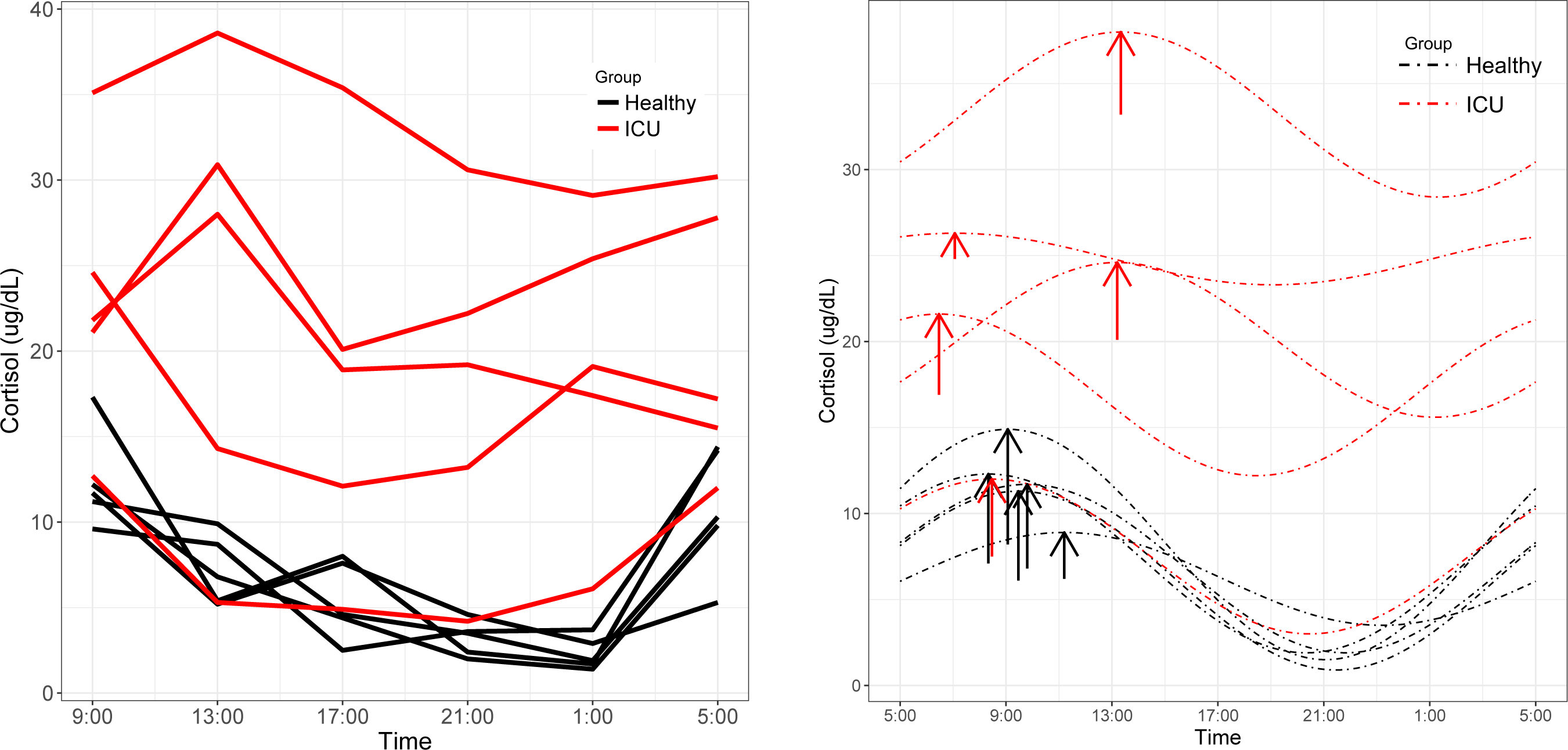
Cortisol rhythms. Left: Individual time series plots of cortisol concentrations quantified by Fairview Hospital Laboratories. Red lines indicate measured cortisol concentrations in ICU patients and black lines indicate healthy controls. Right: ICU patients showed alterations in MESOR and phase not present in controls. Dashed lines represent cosinor fits to the data. Arrows are placed at each rhythm’s acrophase. Arrow lengths reflect rhythm amplitude.

For the 10 putatively identified rhythmic metabolites, some patterns were evident in the data. For cortisone, Lyso PC 18.2, Lyso PE 18.1, Lyso PE 18.2, and proline, rhythms exist both at the individual level and the group level in healthy controls. Rhythms do not exist in ICU patients at either among individuals or as a group, except proline has a rhythm at the individual level in ICU patients.

For acetylcarnitine, aminoadipate, tyramine, and 2-hydroxylauroylcarnitine, neither patients nor controls have significant rhythms when evaluated individually. We interpret this to mean that patients did not differ much from healthy controls individually in these variables. However, PMC showed the healthy control group had significant rhythms while ICU patient group did not. This implies that among ICU patients, there was more variability among individuals in amplitude, MESOR, and acrophase. Among healthy controls, however, there was much less variability among individuals in 24-hour rhythm characteristics.

### Principal component analysis

To more globally assess differences in metabolism between healthy controls and ICU patients, we performed principal component analysis (PCA) on the mass spectrometry data. A scores plot (Fig 4) of the features identified by Progenesis QI shows that each ICU patient’s plasma samples (P07-P12) are strongly clustered together away from healthy controls (P01-P06). Of the 5 ICU patients studied, 4 had ICU stays of >20 days and 3 were discharged to long-term acute care facilities (Table 1). The 5th patient had an 8-day stay and was discharged home. This patient’s cortisol rhythm was closest to the rhythms of healthy controls (Fig 3) and their metabolic profiles (red circles) closest to the cluster of healthy controls (Fig 4).

**Fig 4:**
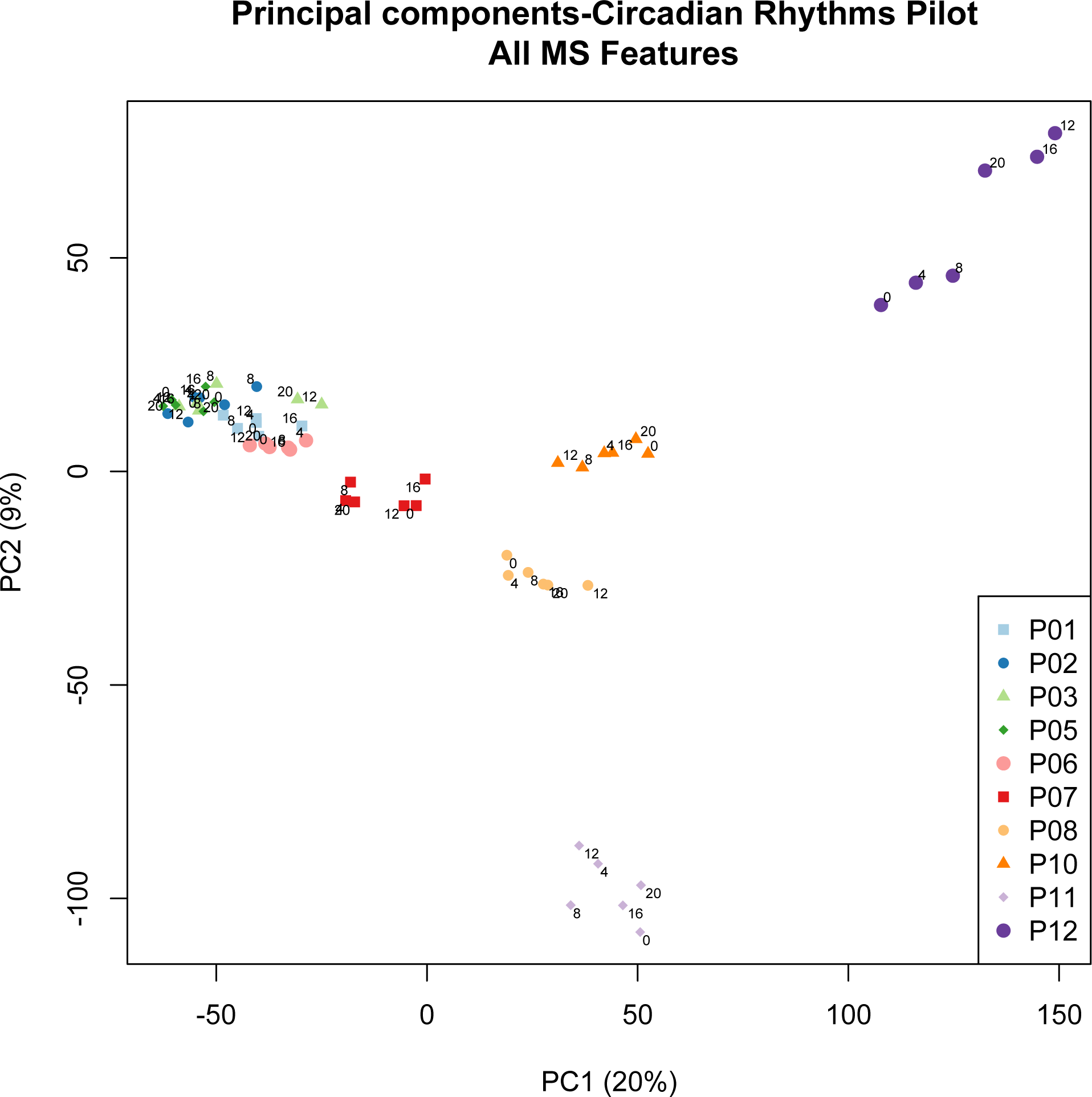
PCA scores plot of metabolic profiles. Scores from the first two principal components were constructed from a total of 60 plasma samples. Samples were obtained from 5 healthy controls (P01-P06, clustered on the left hand side) and 5 ICU patients (P07-P12). Each colored dot corresponds to an individual plasma sample. Each ICU patient is clearly visible in the PCA scores (dark red, orange, yellow, light purple, dark purple). Small numbers indicate the hour at which the plasma sample was obtained (0hr, 4 hr, 8hr, 12hr, 16hr, 20hr). 0hr corresponds to 9:00 AM for all participants. © 2019 Lusczek ER. Published in Metabolomics - New Insights into Biology and Medicine under CC BY 3.0 license. Available from: DOI: 10.5772/intechopen.87145.

To reduce this dataset, features were evaluated for statistical significance in Progenesis QI by comparing feature intensity in the ICU group vs. feature intensity in the healthy control group. Features with ANOVA q<0.05 were retained. This reduced the data set from 25,000 identified features to15,000 features. Since this is still a very large dataset, we explored it with Ingenuity Pathway Analysis (IPA). The canonical pathway analysis showed 17 pathways dysregulated in ICU patients relative to healthy controls at the level of p<0.05 (S6 Table). The top 5 dysregulated pathways were eicosanoid signaling, prostanoid biosynthesis, FXR/RXR activation, spermine biosynthesis, and spermidine biosynthesis I. A striking 68 different diseases and functions were identified by IPA at the level of p<0.05. The top 5 diseases identified were inflammatory response and disease, neurological disease, immunological disease, organismal injury and abnormalities, and cell death and survival (S7 Table).

## Discussion

In this pilot study of 5 ICU patients and 5 healthy controls, we postulated and confirmed that patients’ 24-hour rhythms differ from those of healthy controls. Rhythms were monitored in heart rate, blood pressure, temperature, and circulating plasma metabolites including cortisol. We hypothesized that fewer of these variables would show significant 24-hour rhythms in the ICU patient group than in the healthy control group. Our results support this hypothesis. Healthy controls generally had significant 24-hour rhythms individually and as a group. On the other hand, while a few ICU patients had significant 24-hour rhythms in isolated variables, no significant rhythms were identified in ICU patients at the group level, except in cortisol. This indicates a lack of coherence in rhythm phases and amplitudes among ICU patients. The general lack of a significant rhythm in ICU patients at the population level suggests that 24-hour rhythms are synchronized in healthy controls only.

### Sound levels

Noise levels were significantly louder in ICU patient rooms than in healthy control rooms at all times, and remained elevated at night. In contrast, healthy controls’ rooms had a clear decrease in nighttime noise levels.

The World Health Organization recommends that patient care areas not exceed 35 dB LA_eq_ (36). In this study, we observed LA_eq_ levels far above this threshold in ICU patient rooms. Other groups have observed similar trends (15, 37, 38). It has been known for over 20 years that high noise levels are present in the ICU and that they adversely affect sleep (15, 39, 40). There is a clear link between sleep deprivation and delirium, which is related to poor outcomes in ICU patients (11). Because of this link, protocols to improve sleep in the ICU have been well-described (41, 42). However, these interventions do not explicitly control sound levels, nor is it standard clinical practice to control light or sound levels in the ICU. Simple interventions to control ICU light and sound levels and foster sleep are promising, and show a reduction in the incidence of delirium (43).

Though study limitations prevent us from reporting on light levels in study participants’ rooms, it is worth commenting on since light is a strong synchronizer of the circadian system. Interestingly, some groups have observed a lack of bright light levels as well as a lack of 24-hour variation in light levels in ICU patient rooms, which may contribute to circadian rhythm disruption in patients (16, 17, 44). Protocols to enforce strict light-dark cycles with sufficient brightness in ICU patient rooms during the day should be standard-of-care to help synchronize patient circadian rhythms.

### 24-hour rhythms

Healthy controls had statistically significant or near-significant 24-hour rhythm in heart rate, blood pressure, and temperature at both the individual level and the population level. ICU patients did not have significant 24-hour rhythms in any of the vitals measured. This finding reflects a disruption to 24-hour rhythms in ICU patients, which other groups have noted (6, 45).

We made preliminary identifications of 60 circadian metabolites in the plasma of study participants using mass spectrometry, 10 of which had a significant 24-hour rhythm in the healthy control group. With the exception of cortisol, none of these metabolites showed a significant rhythm in ICU patients at the group level. Even though both ICU patients and healthy controls showed a significant 24-hour rhythm in laboratory-quantified cortisol, we noted that patients’ cortisol rhythms showed abnormalities in phase and MESOR. Elevated cortisol and disrupted cortisol metabolism are known to occur in critical illness (46, 47). Interestingly, the ICU patient with the lowest APACHE II score has a cortisol pattern that is very close to that of the healthy controls.

It is striking that even in this pilot study, profiled physiological indicators of circadian rhythms showed a significant or near-significant rhythm in healthy controls but not in ICU patients. The data identify a significant, global disruption to the circadian rhythm in critical illness. The implications of this finding mandate further study.

### Patient variability

Our work uncovered a surprising clustering pattern in plasma metabolic profiles. Samples from ICU patients formed very distinct clusters corresponding to each individual patient, while samples from healthy controls clustered tightly together, with no resolution at the level of the individuals providing the samples. The ICU patient with plasma samples clustered nearest to the group of healthy controls (red dots in Fig 4) had a similar cortisol rhythm, a lower APACHE II score, and a shorter ICU stay than the other patients. Therefore, we hypothesize that the patient-level resolution of the metabolomics data shown in Fig 4 is linked to an individual patient’s state, and that multiple-day sampling could show patient “trajectories” that move toward improved or worsening health. However, we could discern no other pattern in the PCA scores in this small study. While other researchers have associated metabolomics data with mortality in critical illness (48), our finding raises the possibility that metabolomics may provide broad insight into individual patients’ conditions. More focused research will have to be done to confirm and explain the highly individualized patterns in sample clustering observed in ICU patients.

### Disruptions to metabolism

Restricting the untargeted metabolomics data from 25,000 features to those that differ between ICU patients and healthy controls at the level of q<0.05 only reduced the data set to 15,000 features. These results indicate that ICU patients have highly disordered metabolism relative to healthy controls.

To explore this, we analyzed the untargeted metabolomics data with Ingenuity Pathway Analysis. Interestingly, each of the top 5 dysregulated pathways identified by IPA have been linked to the circadian system. Eicosanoids are broadly involved in inflammation, vasoconstriction, pain perception, and cell growth and regulation. Prostanoids, a class of eicosanoids, have long been known to oscillate with a daily rhythm in saliva (49). The FXR/RXR activation pathway modulates bile acid, lipid, and glucose metabolism. Bile acid synthesis, FXR, and RXR are all linked by the circadian system (50, 51). Finally, spermines and spermidines belong to the polyamide class of molecules and regulate various genetic processes. Polyamine biosynthesis has also been shown to intersect with the circadian system (52).

This pilot study had several limitations. We did not use a circadian protocol with healthy controls or ICU patients. As such, we are unable to clearly state whether the observed differences in 24-hour rhythms are due to endogenous factors, exogenous factors, or both. This warrants further study. Only 5 individuals per group were studied. Future work must include larger groups. All 5 ICU patients had different underlying conditions. A more focused study of patient groups with distinct underlying conditions (e.g. sepsis *vs*. stroke) should be done to evaluate how individual diseases contribute to circadian rhythm desynchronization and to observed patterns in the metabolomics data. Core body temperature should be measured in all participants. The lack of significant rhythm in temperature observed in healthy controls (p=0.091) may be due to this.

Due to budgetary and logistical constraints, only positive mode reverse phase MS1 data were collected. Future work should include negative mode reverse phase and HILIC columns as well as MS2 data to aid in more complete feature identification. The feature identification reported here is preliminary and should be confirmed by targeted analyses in future work. Melatonin was not conclusively identified in our mass spectrometry data and should be quantified by mass spectrometry and/or radio immunoassay in future work. The IPA results should be considered preliminary as well. Progenesis QI can assign more than one HMDB ID to a feature, and IPA resolves duplicates by choosing the feature with the lowest q-value. This may introduce biasing.

More frequent sampling of blood for metabolomics data would improve profiling of circadian rhythms (53). The lack of statistical significance in individual rhythms of metabolites, particularly in healthy controls, may be alleviated by more data. Plasma samples could be collected at a higher frequency, or for a longer period of time, or both. This supports our case that a larger study is needed.

Finally, we recorded bedside light levels for all study participants. However, our light meter had to be replaced halfway through the study and we did not feel that the data from the two separate meters could be meaningfully compared. We have opted to not present these data in the manuscript. Despite these limitations, the data clearly show a global disruption to 24-hour rhythms in ICU patients as well as fascinating individual patterns in ICU patients’ metabolomes. Both results deserve further study.

## Conclusions

This pilot study confirms the literature supporting a broad desynchronization of circadian rhythms in ICU patients on multiple levels by measuring circadian rhythms in vital signs and circulating metabolites. In addition, PCA of untargeted metabolomics identifies a strong pattern in metabolomes of ICU patients, which are highly individualized and distinct from healthy controls. These results suggest a significant disordering of physiology in ICU patients, involving at a minimum the metabolism and the circadian system, which should be characterized further in order to determine the implications for patient care.

## Data Availability

Data are available at the Data Repository for the University of Minnesota (DRUM).

https://conservancy.umn.edu/handle/11299/208811

## Acknowledgements

The authors would like to thank Julie Kirihara PhD and the Center for Mass Spectrometry and Proteomics at the University of Minnesota for assistance in obtaining mass spectrometry data, and the University of Minnesota Medical Center ICU staff.

## Author contributions

- ERL was study PI. She contributed to study design, data analysis and interpretation, and prepared the manuscript.
- JE and LP performed data analysis and assisted with manuscript preparation and approved the final manuscript.
- SH obtained mass spectrometry data and contributed to manuscript preparation and approved the final manuscript.
- MS and SM identified patients and collected data, and approved the final manuscript.
- GB contributed to study design and data interpretation, and approved the final manuscript.
- GC-G contributed to study design, data analysis and interpretation, manuscript preparation, and approved the final manuscript.

## Supporting Information

S1: Deuterated internal standards used in sample preparation. Standards were obtained from CDN Isotopes and Cambridge Isotope Laboratories.

S2: BMI data for health controls and ICU patients.

S3: Time series plots of vitals and circadian features putatively identified with mass spectrometry. Healthy controls are shown in black and ICU patients are shown in red.

S4: Time series plots of circadian features putatively identified with mass spectrometry. Spectral intensities are plotted against time as mean with standard error bars. Cosinor fits of the 24-hour rhythm are shown in dotted lines. Red indicates ICU patients and black indicates healthy controls. Two features were ambiguously assigned the identity Lyso PE 18.2 and both are shown.

S5: Cortisol spectral intensities quantified with mass spectrometry. Overall there is good agreement with the laboratory-quantified cortisol concentrations.

S6: Canonical pathways dysregulated in ICU patients relative to healthy controls. Pathways were identified by Ingenuity Pathway Analysis of the mass spectrometry data set.

S7: Diseases and Functions in ICU patients relative to healthy controls. Diseases and functions were identified by Ingenuity Pathway Analysis of the mass spectrometry data set.

**Supplemental Table 1:** BMI Data.

| **Patient ID**<br>**(Healthy)** | **BMI** | **Patient ID**<br>**(ICU)** | **BMI** |
| --- | --- | --- | --- |
| P01 | 31.6 | P07 | 31.0 |
| P02 | 22.5 | P08 | 37.1 |
| P03 | 22.4 | P10 | 35.1 |
| P05 | 30.3 | P11 | 37.2 |
| P06 | 33.3 | P12 | 57.3 |

BMI data for healthy controls and ICU patients. At the group level, BMI did not significantly differ between healthy controls and ICU patients ( $p=0.07$ ).

Supplemental Files 6 and 7 available upon request

**Supplemental Table 2:** Deuterated internal standards.

| Compound | MW | +H | -H | conc. |
| --- | --- | --- | --- | --- |
| Phenylalanine-D5 | 170.10981 | 171.11764 | 169.10179 | 50ug/mL |
| Hippuric Acid-D5 | 184.08908 | 185.0969 | 183.08111 | 50ug/mL |
| Cholic acid-D4 | 412.31213 | 413.31996 | 411.3054 | 50ug/mL |
| Glucose-D7 | 187.10678 | 188.1146 | 186.10005 | 500ug/mL |
|  |  | 210.09655(Na) | 222.0762 (Cl) |  |
| C16:0-D3 (palmitic acid) |  |  | 258.25138 | 20ug/mL |

Deuterated standards were obtained from CDN Isotopes and Cambridge isotopes Laboratories.

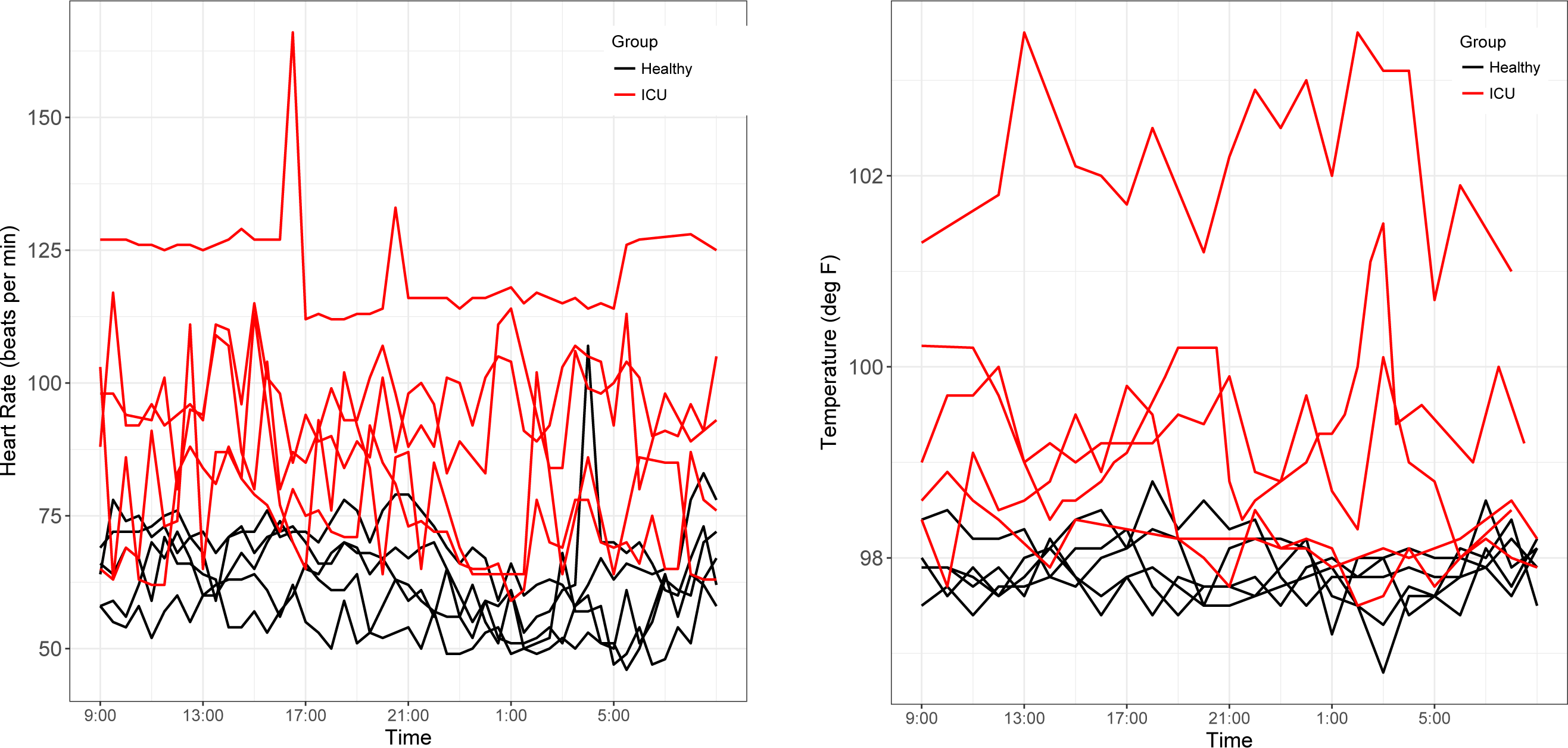

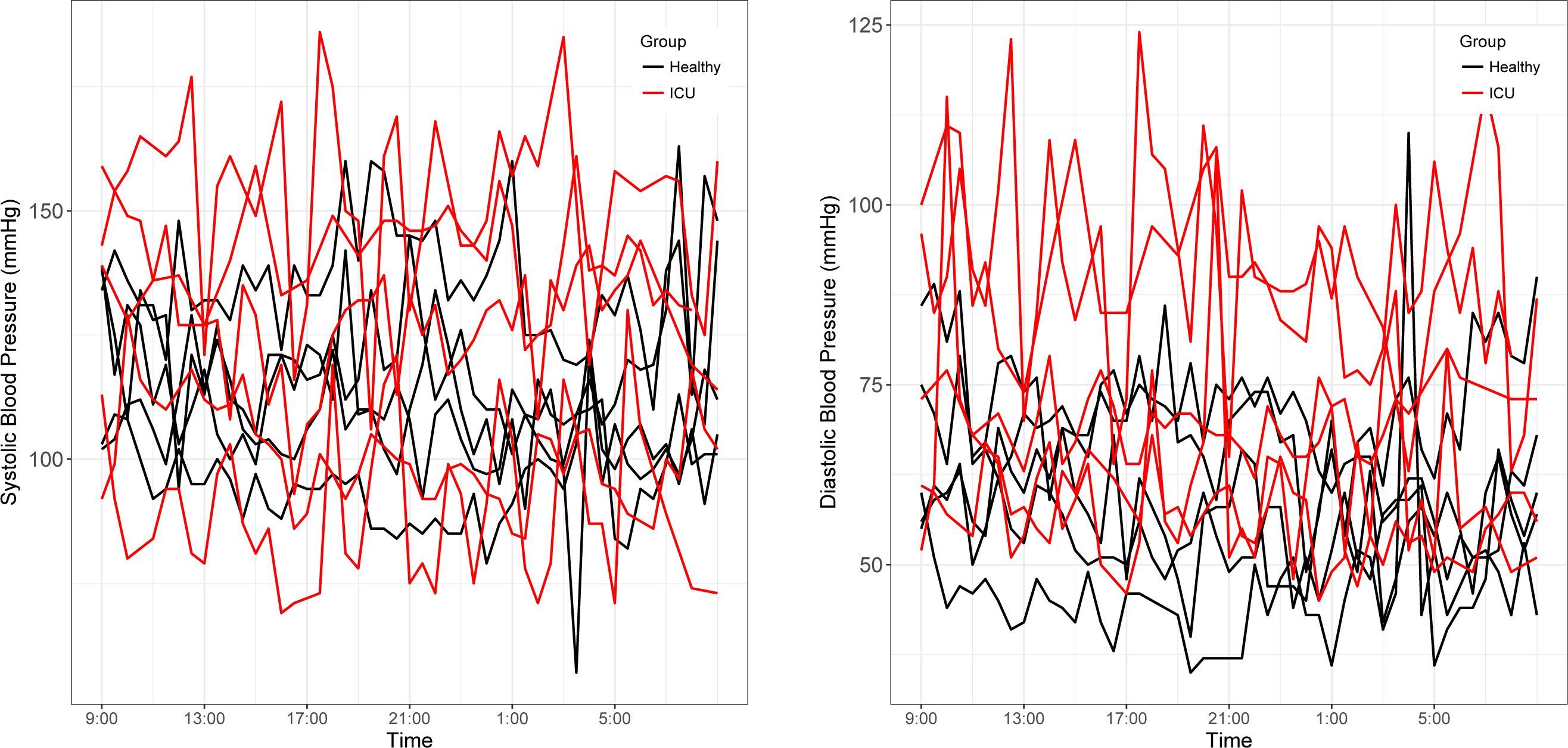

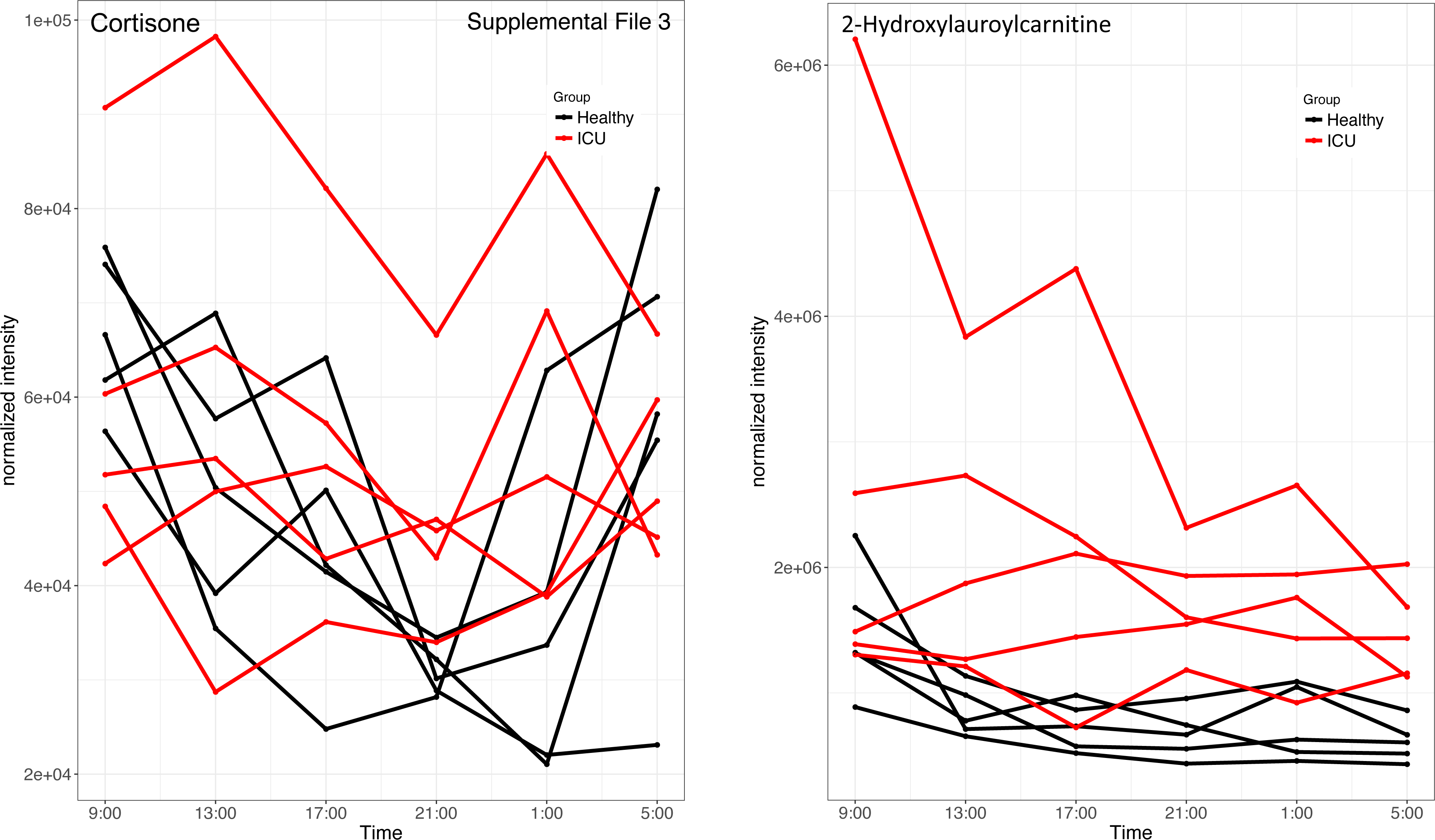

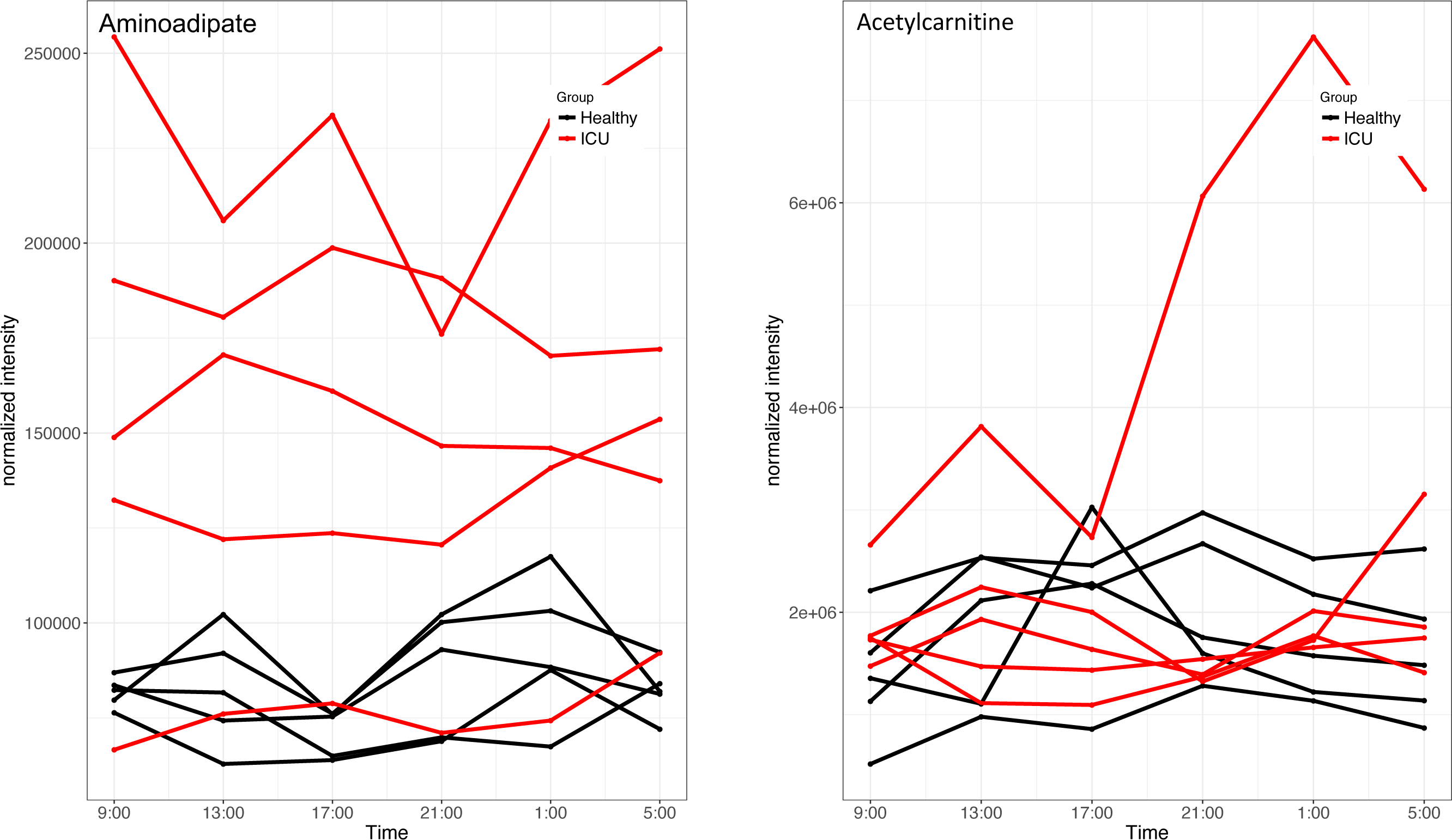

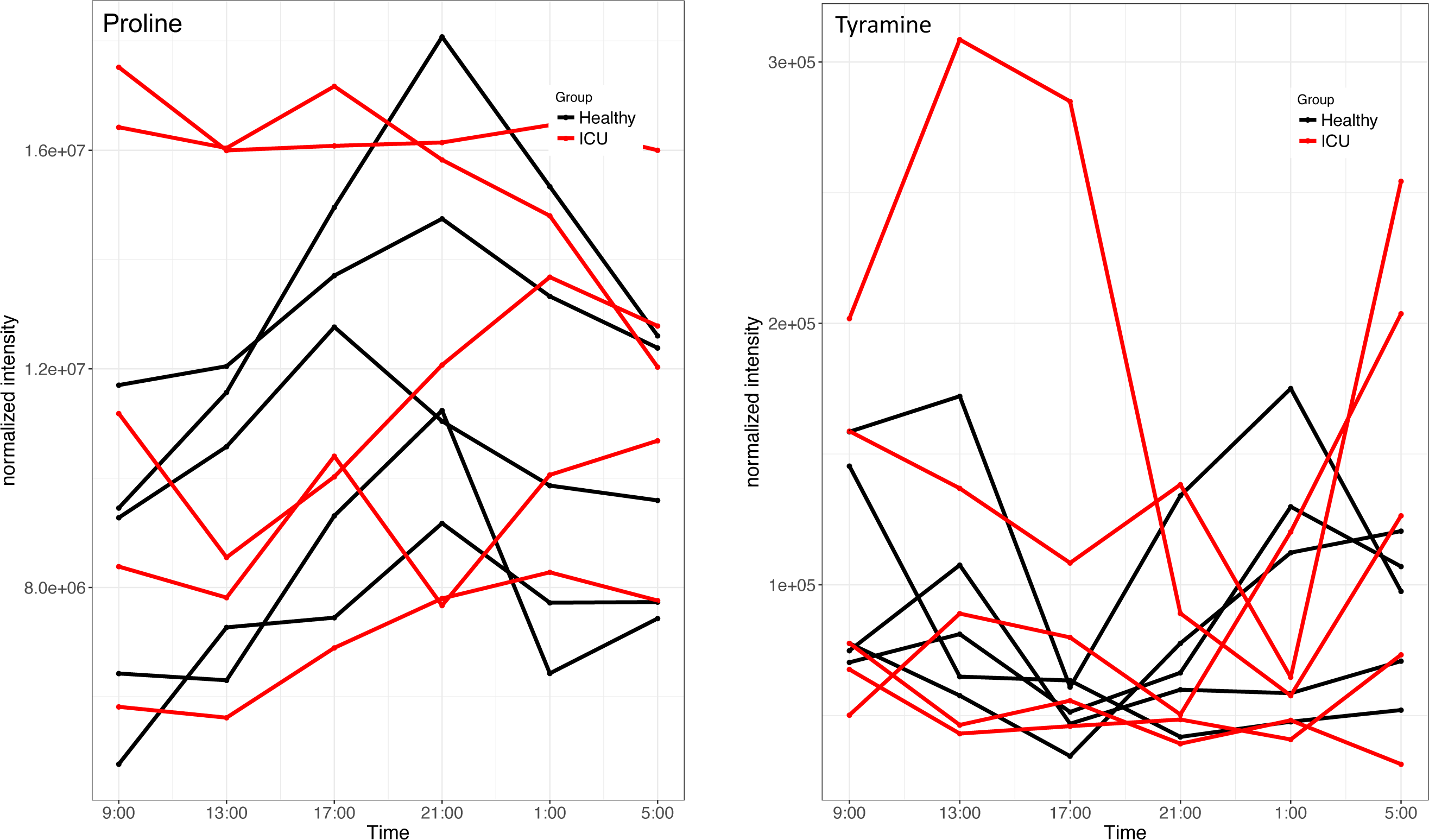

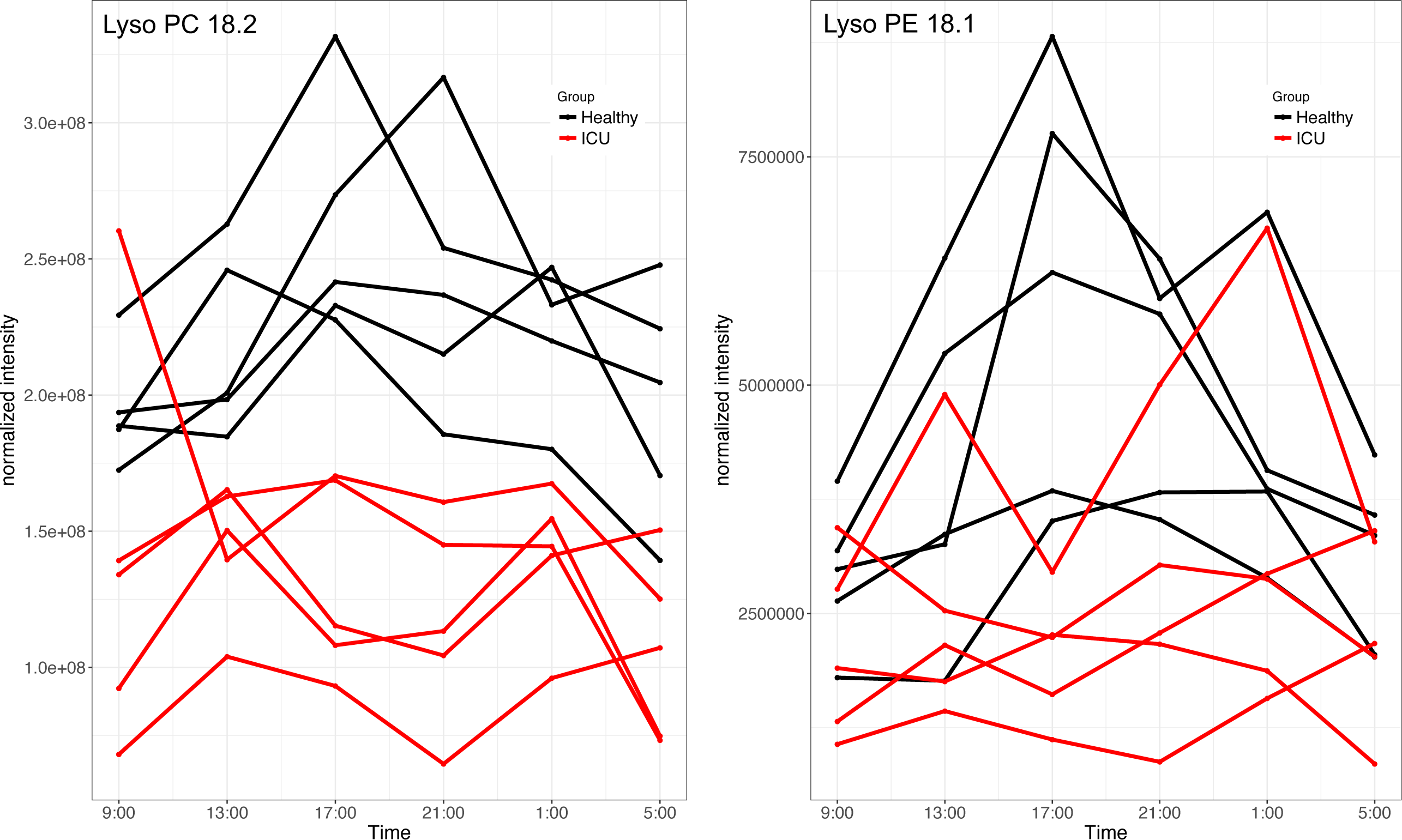

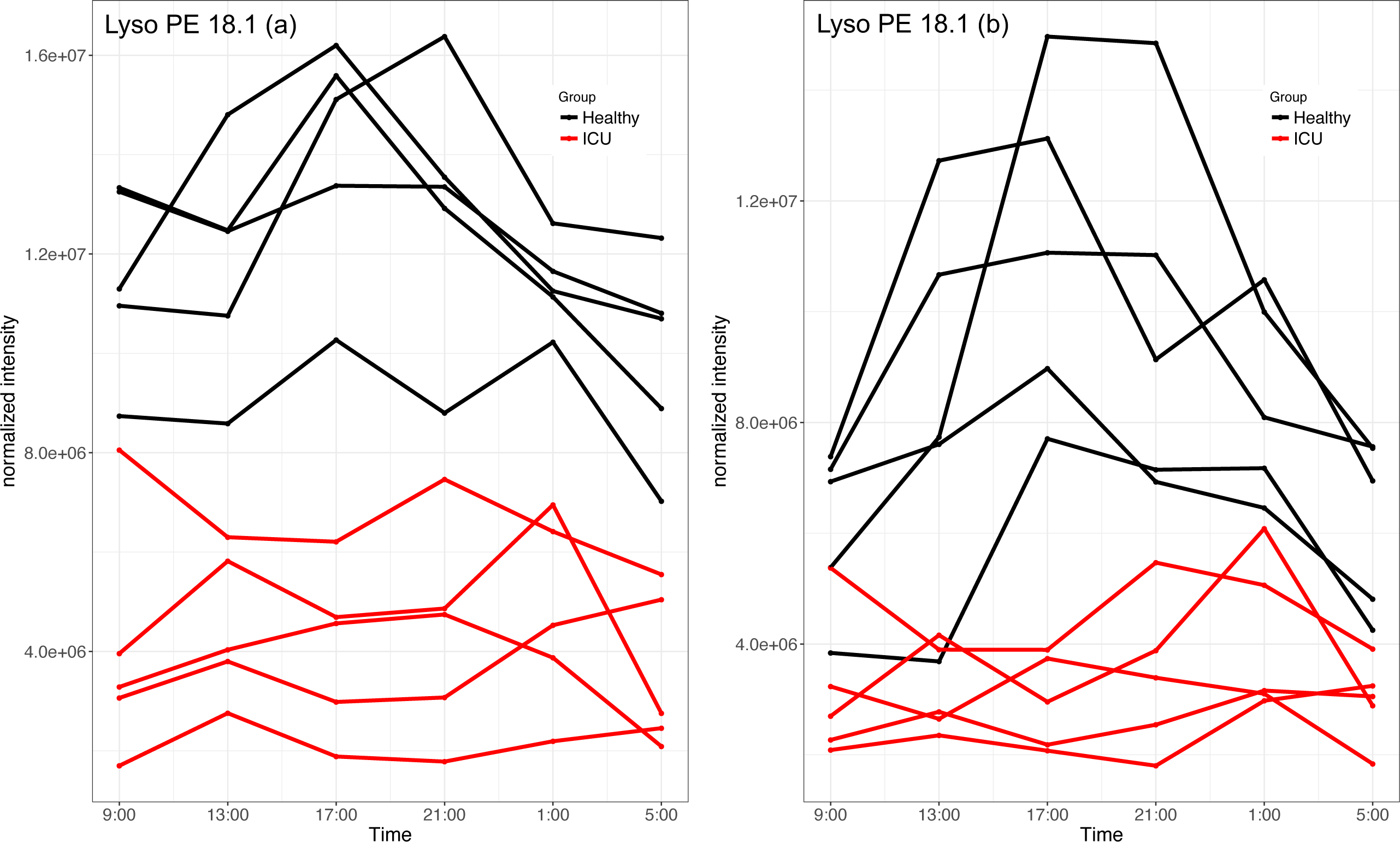

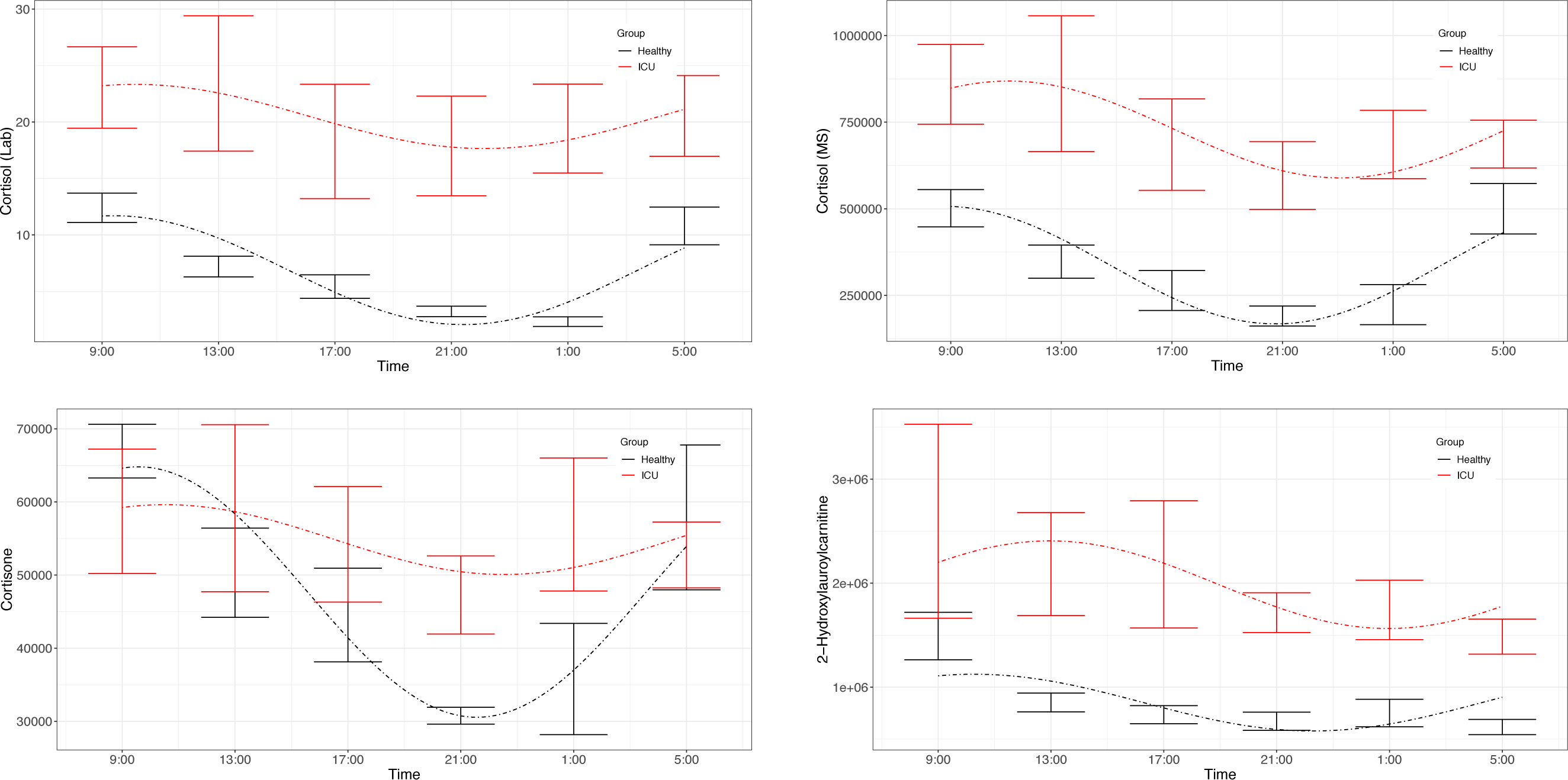

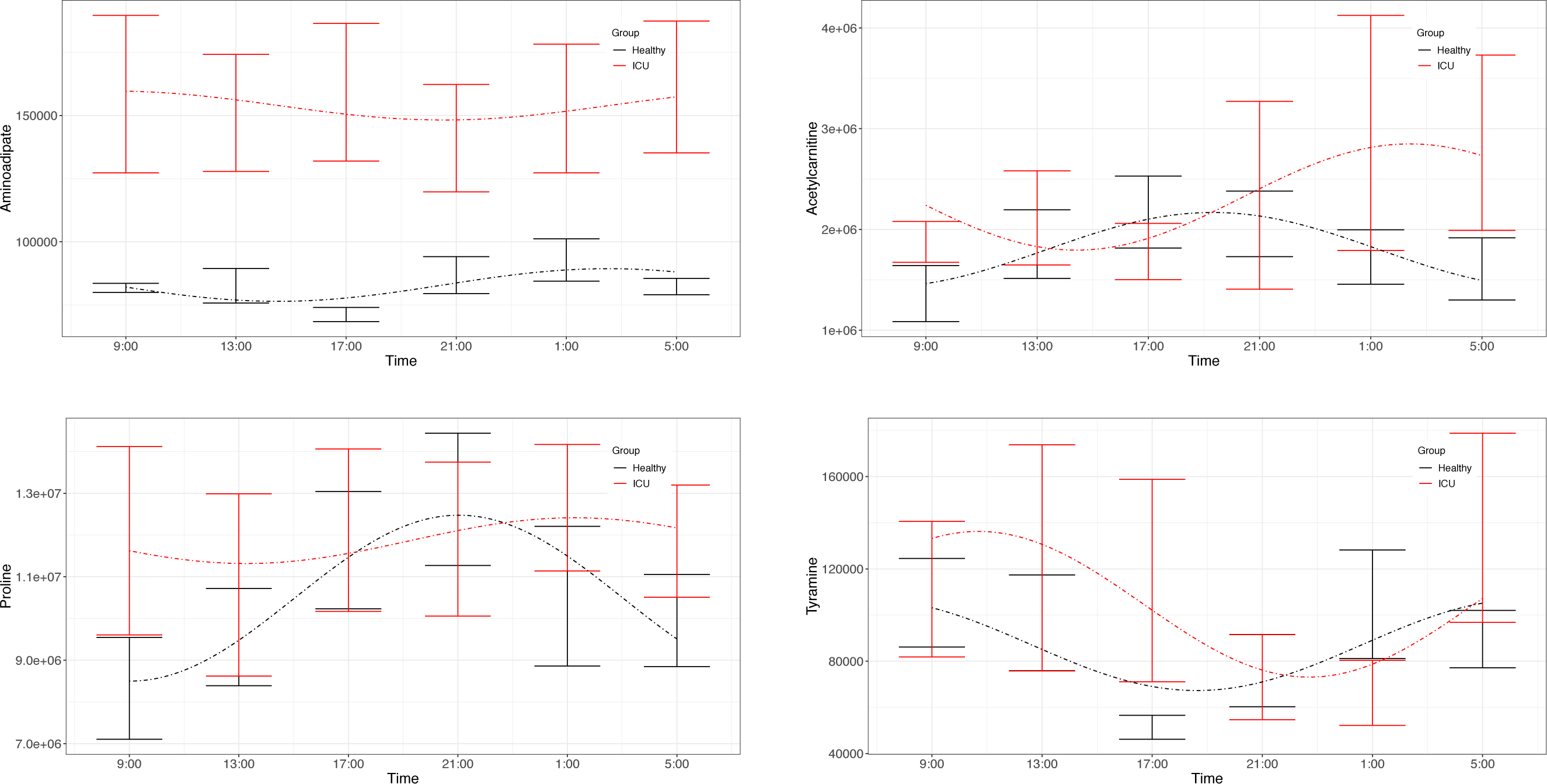

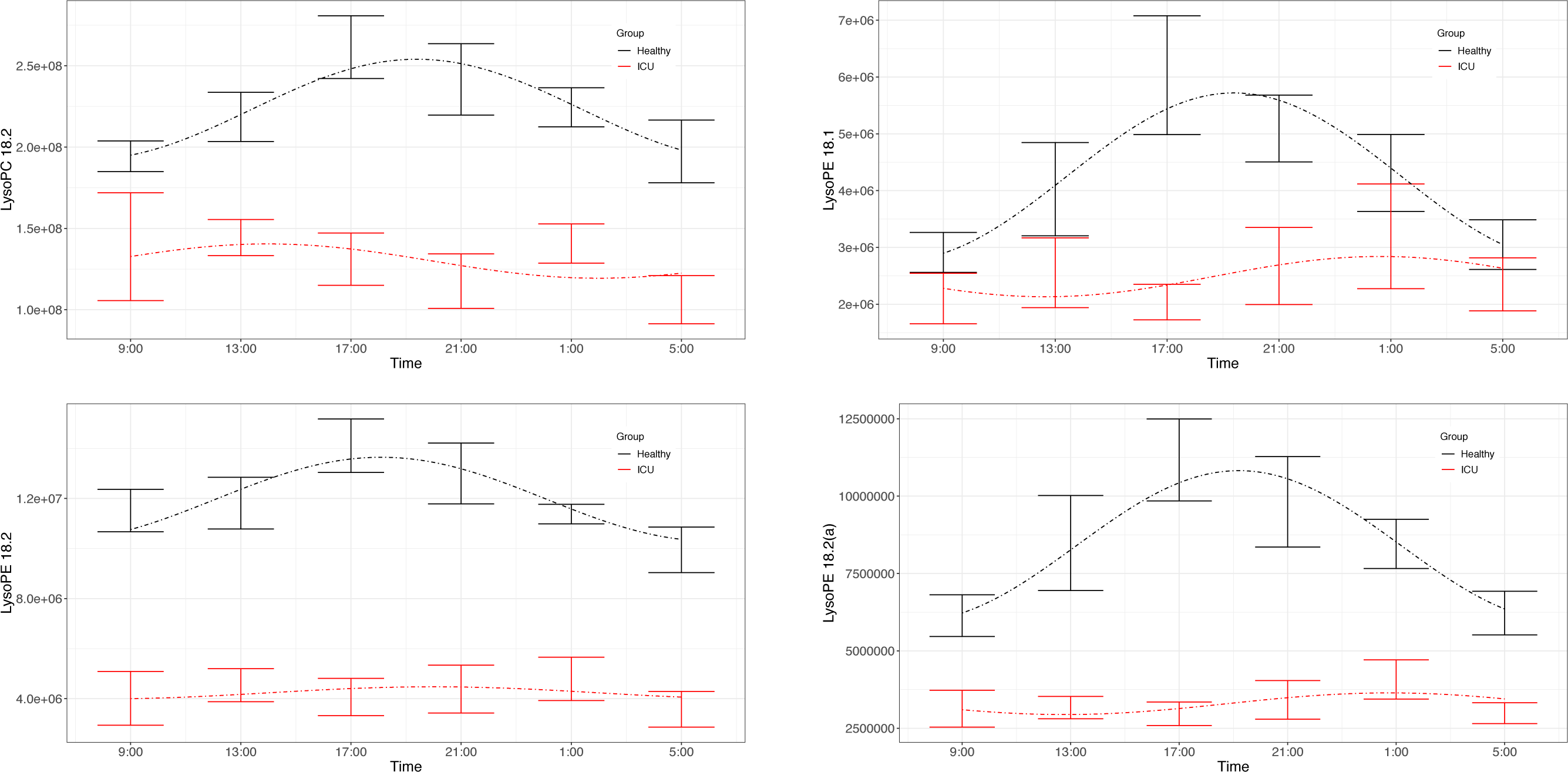

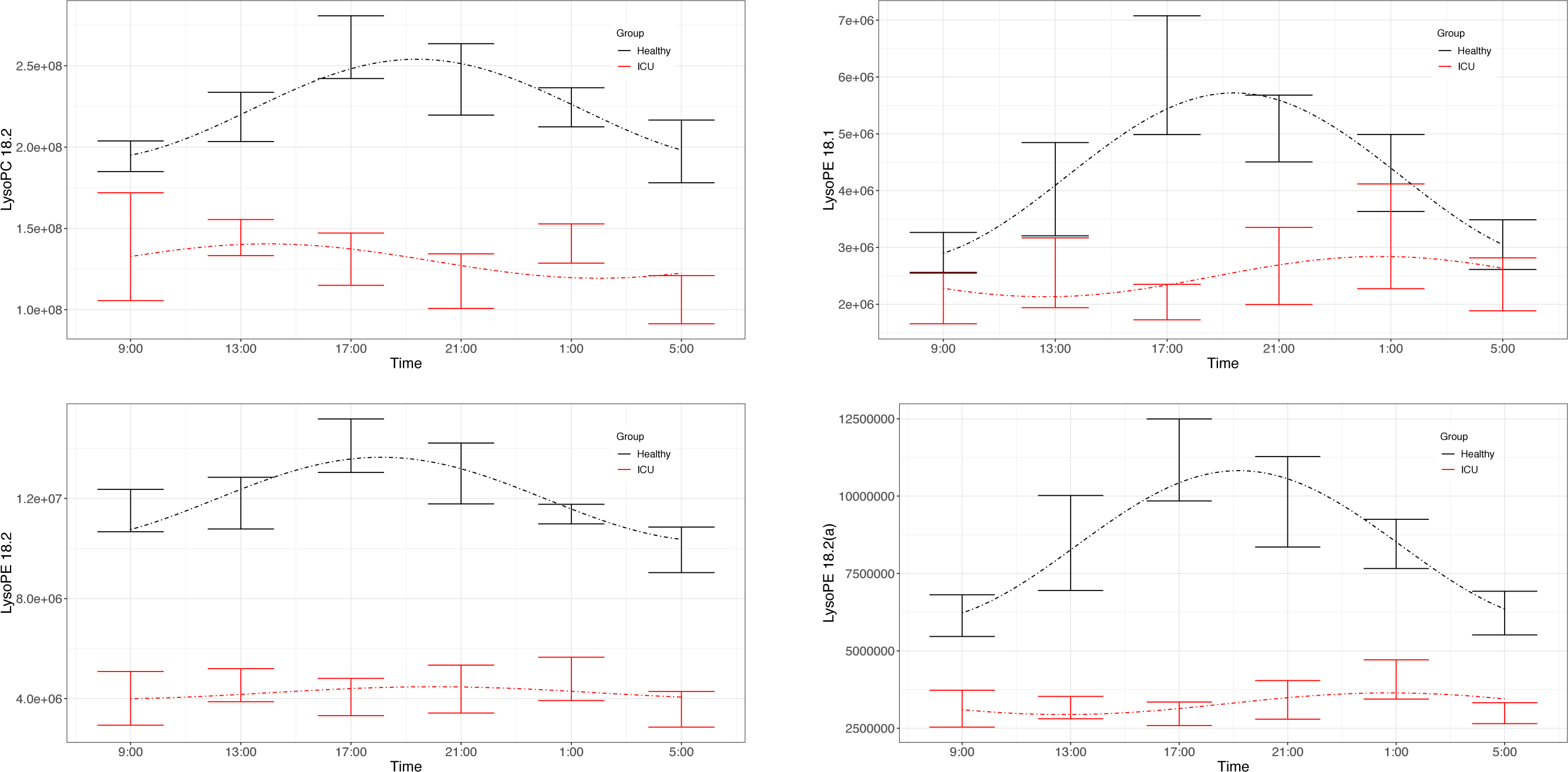

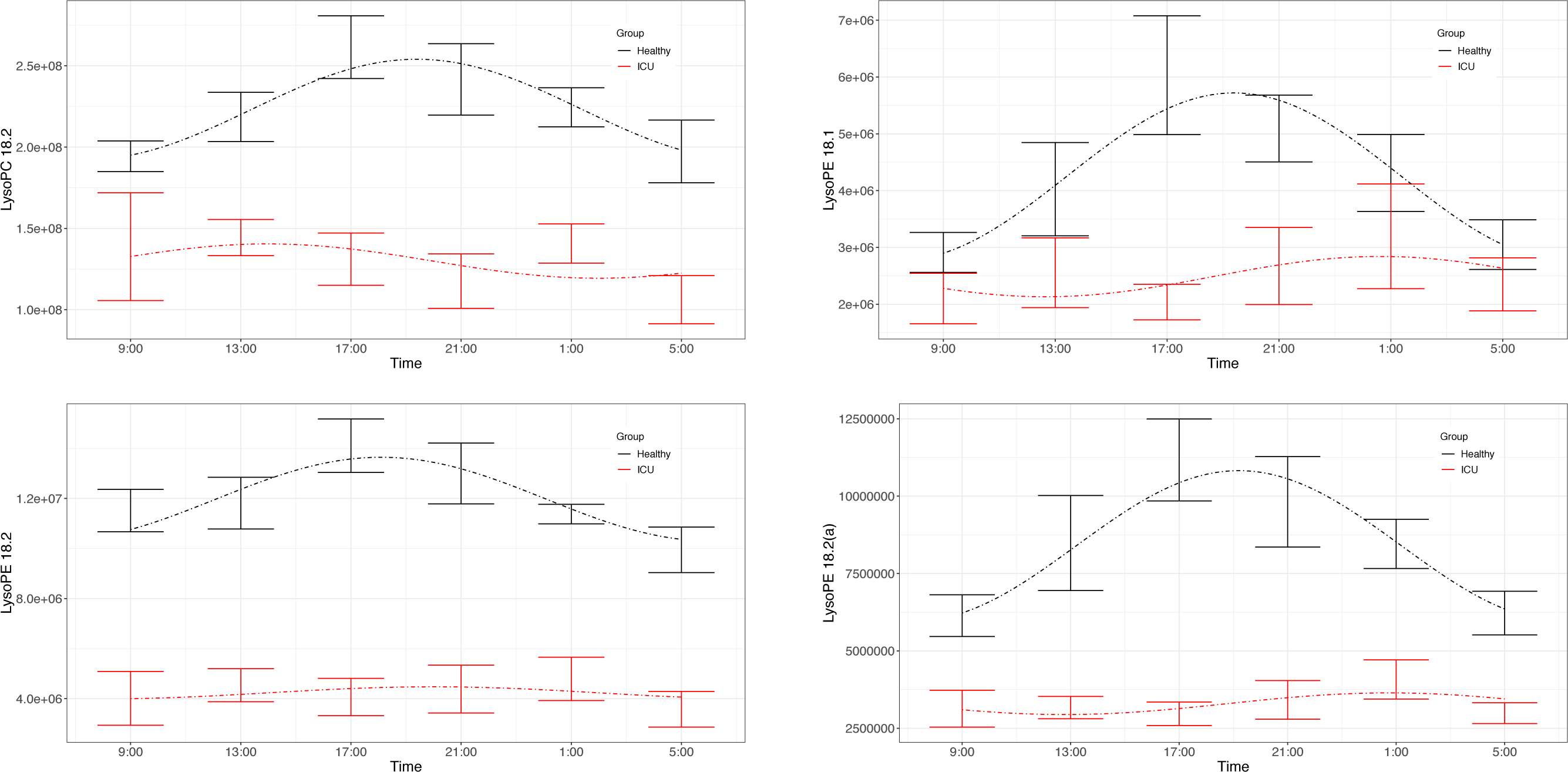

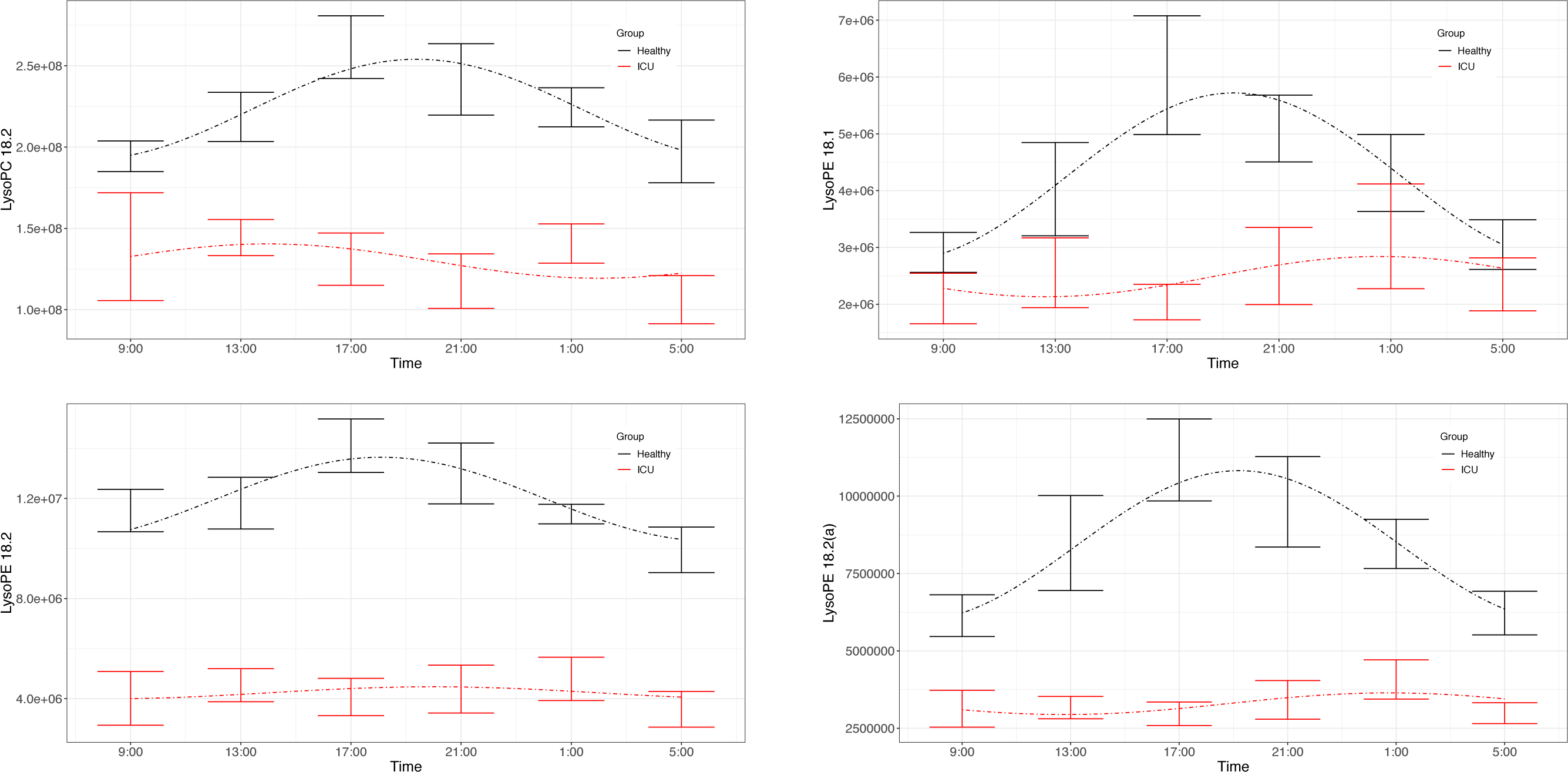

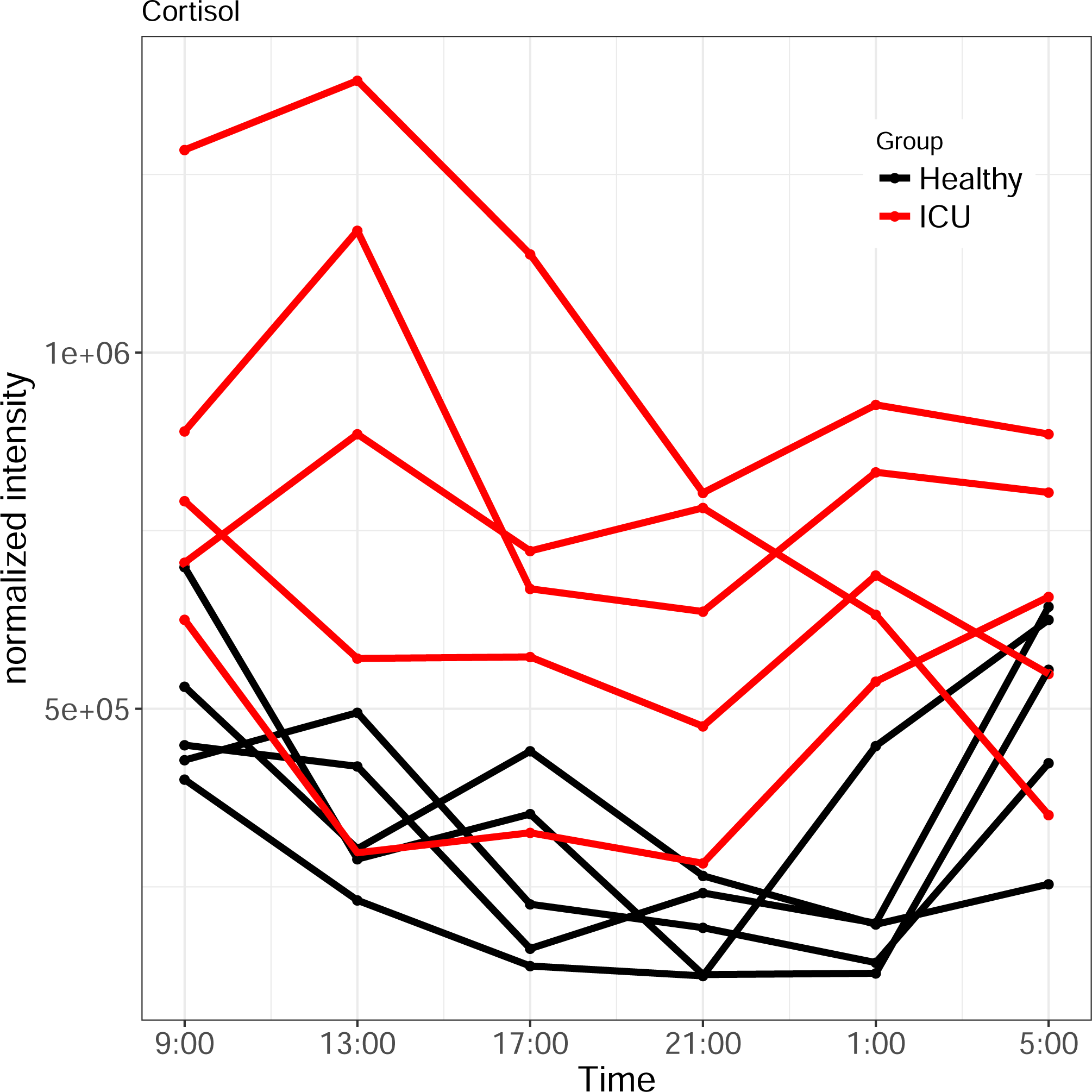

## Notes

### Competing Interest Statement

The authors have declared no competing interest.

### Clinical Trial

This is not a clinical trial.

### Funding Statement

Funding was provided by the Department of Surgery at the University of Minnesota [ERL] and by the Halberg Chronobiology Fund [GC-G].

## References

1. Mohawk JA, Green CB, Takahashi JS. Central and peripheral circadian clocks in mammals. Annual Review of Neuroscience. 2012;35:445–62.

2. Turek FW. Circadian clocks: Not your grandfather’s clock. Science (New York, NY). 2016;354(6315):992–3.

3. Bass J, Lazar MA. Circadian time signatures of fitness and disease. Science. 2016;354(6315):994–9.

4. Roenneberg T, Merrow M. The Circadian Clock and Human Health. Current Biology. 2016;26(10):R432–R43.

5. McKenna HT, Reiss IK, Martin DS. The significance of circadian rhythms and dysrhythmias in critical illness. Journal of the Intensive Care Society. 2017;18(2):121–9.

6. Chan M-C, Spieth PM, Quinn K, Parotto M, Zhang H, Slutsky AS. Circadian rhythms: From basic mechanisms to the intensive care unit *. Critical Care Medicine. 2012;40(1):246–53.

7. Hrushesky W, Grutsch J, Huff DF, Kazlausky T, Tavolacci L. Temporal tuning of daily rhythms helps advanced cancer patients and cancer survivors feel better, live better, and live longer. ChronoPhysiology and Therapy. 2016;6:1–13.

8. Oldham MA, Lee HB, Desan PH. Circadian Rhythm Disruption in the Critically Ill: An Opportunity for Improving Outcomes. Critical Care Medicine. 2016;44(1):207–17.

9. McKenna H, van der Horst GT, Reiss I, Martin D. Clinical chronobiology: a timely consideration in critical care medicine. Critical Care. 2018;22(1):124.

10. Knauert MP, Haspel JA, Pisani MA. Sleep loss and circadian rhythm disruption in the intensive care unit. Clinics in chest medicine. 2015;36(3):419–29.

11. Figueroa-Ramos MI, Arroyo-Novoa CM, Lee KA, Padilla G, Puntillo KA. Sleep and delirium in ICU patients: a review of mechanisms and manifestations. Intensive care medicine. 2009;35(5):781–95.

12. Miki H, Yano M, Iwanaga H, Tsujinaka T, Nakayama M, Kobayashi M, et al. Total parenteral nutrition entrains the central and peripheral circadian clocks. Neuroreport. 2003;14(11):1457–61.

13. Matuchansky C, Messing B, Beliah M, Jeejeebhoy K, Beau P, Allard J. Cyclical parenteral nutrition. The Lancet. 1992;340(8819):588–92.

14. Rothschild J, Lagakos W. Implications of enteral and parenteral feeding times: considering a circadian picture. Journal of Parenteral and Enteral Nutrition. 2015;39(3):266–70.

15. Elbaz M, Léger D, Sauvet F, Champigneulle B, Rio S, Strauss M, et al. Sound level intensity severely disrupts sleep in ventilated ICU patients throughout a 24-h period: a preliminary 24-h study of sleep stages and associated sound levels. Annals of intensive care. 2017;7(1):25.

16. Danielson SJ, Rappaport CA, Loher MK, Gehlbach BK. Looking for light in the din: An examination of the circadian-disrupting properties of a medical intensive care unit. Intensive and Critical Care Nursing. 2018.

17. Durrington HJ, Clark R, Greer R, Martial FP, Blaikley J, Dark P, et al. ‘In a dark place, we find ourselves’: light intensity in critical care units. Intensive care medicine experimental. 2017;5(1):9.

18. Zhang R, Lahens NF, Ballance HI, Hughes ME, Hogenesch JB. A circadian gene expression atlas in mammals: implications for biology and medicine. Proceedings of the National Academy of Sciences of the United States of America. 2014;111(45):16219–24.

19. Asher G, Schibler U. Crosstalk between components of circadian and metabolic cycles in mammals. Cell Metabolism. 2011;13(2):125–37.

20. Bass J, Takahashi JS. Circadian integration of metabolism and energetics. Science. 2010;330(6009):1349–54.

21. Eckel-Mahan KL, Patel VR, Mohney RP, Vignola KS, Baldi P, Sassone-Corsi P. Coordination of the transcriptome and metabolome by the circadian clock. Proceedings of the National Academy of Sciences. 2012;109(14):5541–6.

22. Skene DJ, Skornyakov E, Chowdhury NR, Gajula RP, Middleton B, Satterfield BC, et al. Separation of circadian-and behavior-driven metabolite rhythms in humans provides a window on peripheral oscillators and metabolism. Proceedings of the National Academy of Sciences. 2018;115(30):7825–30.

23. Davies SK, Ang JE, Revell VL, Holmes B, Mann A, Robertson FP, et al. Effect of sleep deprivation on the human metabolome. Proceedings of the National Academy of Sciences of the United States of America. 2014;111(29):10761–6.

24. Dallmann R, Viola AU, Tarokh L, Cajochen C, Brown SA. The human circadian metabolome. Proceedings of the National Academy of Sciences. 2012;109(7):2625–9.

25. Kervezee L, Cermakian N, Boivin DB. Individual metabolomic signatures of circadian misalignment during simulated night shifts in humans. PLoS biology. 2019;17(6):e3000303.

26. Honma A, Revell VL, Gunn PJ, Davies SK, Middleton B, Raynaud FI, et al. Effect of Acute Total Sleep Deprivation on Plasma Melatonin, Cortisol and Metabolite Rhythms in Females. European Journal of Neuroscience. 2019.

27. Want EJ, O’Maille G, Smith CA, Brandon TR, Uritboonthai W, Qin C, et al. Solvent-dependent metabolite distribution, clustering, and protein extraction for serum profiling with mass spectrometry. Analytical chemistry. 2006;78(3):743–52.

28. Dunn WB, Broadhurst D, Begley P, Zelena E, Francis-McIntyre S, Anderson N, et al. Procedures for large-scale metabolic profiling of serum and plasma using gas chromatography and liquid chromatography coupled to mass spectrometry. Nature protocols. 2011;6(7):1060.

29. R Core Team (2018). R: A language and environment for statistical computing. R Foundation for Statistical Computing, Vienna, Austria. URL https://www.R-project.org/.

30. Cornelissen G. Cosinor-based rhythmometry. Theoretical Biology and Medical Modelling. 2014;11(1):16.

31. Sokal RR, Rohlf FJ. Biometry, 3rd edn New York. NY: WH Freeman and Company. 1995.

32. Smith CA, O’Maille G, Want EJ, Qin C, Trauger SA, Brandon TR, et al. METLIN: a metabolite mass spectral database. Therapeutic drug monitoring. 2005;27(6):747–51.

33. Wishart DS, Knox C, Guo AC, Eisner R, Young N, Gautam B, et al. HMDB: a knowledgebase for the human metabolome. Nucleic acids research. 2009;37(suppl 1):D603–D10.

34. Ang JE, Revell V, Mann A, Mäntele S, Otway DT, Johnston JD, et al. Identification of human plasma metabolites exhibiting time-of-day variation using an untargeted liquid chromatography–mass spectrometry metabolomic approach. Chronobiology international. 2012;29(7):868–81.

35. Storey JD, Tibshirani R. Statistical significance for genomewide studies. Proceedings of the National Academy of Sciences. 2003;100(16):9440–5.

36. Berglund B, Lindvall T, Schwela DH. Guidelines for community noise: World Health Organization; 1995.

37. Darbyshire JL, Young JD. An investigation of sound levels on intensive care units with reference to the WHO guidelines. Critical Care. 2013;17(5):R187.

38. Knauert M, Jeon S, Murphy TE, Yaggi HK, Pisani MA, Redeker NS. Comparing average levels and peak occurrence of overnight sound in the medical intensive care unit on A-weighted and C-weighted decibel scales. Journal of critical care. 2016;36:1–7.

39. Aaron JN, Carlisle CC, Carskadon MA, Meyer TJ, Hill NS, Millman RP. Environmental noise as a cause of sleep disruption in an intermediate respiratory care unit. Sleep. 1996;19(9):707–10.

40. Meyer TJ, Eveloff SE, Bauer MS, Schwartz WA, Hill NS, Millman RP. Adverse environmental conditions in the respiratory and medical ICU settings. Chest. 1994;105(4):1211–6.

41. Balas MC, Vasilevskis EE, Olsen KM, Schmid KK, Shostrom V, Cohen MZ, et al. Effectiveness and safety of the awakening and breathing coordination, delirium monitoring/management, and early exercise/mobility bundle. Critical Care Medicine. 2014;42(5):1024–36.

42. Barr J, Fraser GL, Puntillo K, Ely EW, Gelinas C, Dasta JF, et al. Clinical practice guidelines for the management of pain, agitation, and delirium in adult patients in the intensive care unit. Critical Care Medicine. 2013;41(1):263–306.

43. Kamdar BB, King LM, Collop NA, Sakamuri S, Colantuoni E, Neufeld KJ, et al. The effect of a quality improvement intervention on perceived sleep quality and cognition in a medical ICU. Critical care medicine. 2013;41(3):800.

44. Fan EP, Abbott SM, Reid KJ, Zee PC, Maas MB. Abnormal environmental light exposure in the intensive care environment. Journal of critical care. 2017;40:11–4.

45. J Madrid-Navarro C, Sánchez-Gálvez R, Martinez-Nicolas A, Marina R, A Garcia J, A Madrid J, et al. Disruption of circadian rhythms and delirium, sleep impairment and sepsis in critically ill patients. Potential therapeutic implications for increased light-dark contrast and melatonin therapy in an ICU environment. Current pharmaceutical design. 2015;21(24):3453–68.

46. Boonen E, Vervenne H, Meersseman P, Andrew R, Mortier L, Declercq PE, et al. Reduced cortisol metabolism during critical illness. New England Journal of Medicine. 2013;368(16):1477–88.

47. Marik PE. Critical illness-related corticosteroid insufficiency. Chest. 2009;135(1):181–93.

48. Rogers AJ, McGeachie M, Baron RM, Gazourian L, Haspel JA, Nakahira K, et al. Metabolomic derangements are associated with mortality in critically ill adult patients. PloS one. 2014;9(1):e87538.

49. Rigas B, Levine L. Human salivary eicosanoids: circadian variation. Biochemical and biophysical research communications. 1983;115(1):201–5.

50. Chiang JY, Pathak P, Liu H, Donepudi A, Ferrell J, Boehme S. Intestinal farnesoid X receptor and Takeda G protein couple receptor 5 signaling in metabolic regulation. Digestive Diseases. 2017;35(3):241–5.

51. De Cosmo S, Mazzoccoli G. Retinoid X receptors intersect the molecular clockwork in the regulation of liver metabolism. Frontiers in endocrinology. 2017;8:24.

52. Zwighaft Z, Aviram R, Shalev M, Rousso-Noori L, Kraut-Cohen J, Golik M, et al. Circadian clock control by polyamine levels through a mechanism that declines with age. Cell metabolism. 2015;22(5):874–85.

53. Hughes ME, Abruzzi KC, Allada R, Anafi R, Arpat AB, Asher G, et al. Guidelines for genome-scale analysis of biological rhythms. Journal of biological rhythms. 2017;32(5):380–93.

